# Childhood food insecurity and youth mental health trajectories in two UK longitudinal cohorts

**DOI:** 10.64898/2025.12.10.25341979

**Authors:** Eileen Y. Xu, Amelia Edmondson-Stait, Alex S.F. Kwong, Heather C. Whalley

## Abstract

**Background:** Food insecurity (FI) is associated with overall poorer mental health in childhood and adolescence and negatively impacts longer-term health. However, less is known about how differential exposure to FI may shape the development of different mental health symptoms over this period.

**Methods:** We used data from two population-based UK birth cohorts: the Avon Longitudinal Study of Parents and Children (ALSPAC; n=6,182; born 1991-1992) and Growing Up in Scotland (GUS; n=3,167; born 2004-2005). FI severity at age ∼5 years (No FI/Low FI/High FI) was determined from parent-reported difficulties affording food. Mental health symptoms were parent-reported over the following 10 years using the conduct, emotional, hyperactivity/inattention and peer problem subscales of the Strengths and Difficulties Questionnaire (SDQ). Trajectories were characterised using multilevel growth curve models adjusted for child sex, household income, maternal mental health and multiple deprivation index.

**Findings:** High FI at age ∼5 years was associated with worse trajectories of conduct, emotional, hyperactivity/inattention and peer problems over the following 10 years in both cohorts (vs. No FI). In ALSPAC, differences in conduct and hyperactivity/inattention scores were most pronounced at ages 7 (0.31 [0.11,0.51], *p_adj_*<.001) and 9 (0.44 [0.09,0.79], *p*_adj_=.008), respectively; these occurred at ages 13 (0.26 [0.04,0.48], *p_adj_*=.014) and 15 (0.43 [0.04,0.81], *p*_adj_=.023) in GUS. For emotional (ALSPAC: 0.46 [0.22,0.69], *p*_adj_<.001; GUS: 0.38 [0.16,0.59], *p*_adj_<.001) and peer problems (ALSPAC: 0.34 [0.13,0.55], p_adj_<.001; GUS: 0.25 [0.05,0.45], p_adj_=.01), differences between High and No FI groups were most pronounced at age 9 instead. Low FI was also associated – to a lesser degree – with higher peer problem trajectories (vs. No FI); this was most pronounced at age 13 in ALSPAC (0.13 [0.00,0.27], *p*=.031) and age 15 in GUS (0.28 [0.04,0.52], *p*=.016).

**Interpretation:** Children who experienced more severe FI had greater levels of mental health symptoms over the next 10 years, even after adjusting for sociodemographic confounders. FI severity may also display a dose-response effect for peer problems. While replication using more robust FI measures remains necessary, we provide further evidence of the persistent negative impact of FI on youth mental health – particularly during late childhood and mid-adolescence.

**Funding:** University of Edinburgh, Wellcome Trust, University of Bristol

**Research in context:** *Evidence before this study:* We searched PubMed for studies examining youth mental health trajectories after early food insecurity (FI) using the following terms: (food) AND (insecur*) AND ((child*) OR (youth) OR (young people) OR (adolescen*)) AND (("mental health") OR (distress) OR (emotion*) OR (behavio*) OR (internali*) OR (externali*)) AND ((trajectory) OR (trajectories)). Three studies investigated the relationship between FI and youth mental health trajectories using data from South Africa, USA and Canada; none investigated the potential impact of marginal food security. Two studies found an association between FI and persistently high depressive and hyperactivity/inattention symptoms over 1-3.5 years of follow-up. The third study used latent growth curve analysis to investigate eight patterns of FI exposure (based on binary FI status over three timepoints) and teacher-reported behaviour problems, reporting no associations. This study, however, only examined linear changes in behavioural problems which typically follow a non-linear trajectory – i.e. age-related differences in rates of change were not considered.

*Added value of this study:* To our knowledge, this is the first study to investigate trajectories of mental health symptoms in two generations of UK children and adolescents following exposure to different levels of FI severity. Across two large, population-based birth cohorts, we found that high FI at age 5 was associated with worse 10-year trajectories of emotional, conduct, hyperactivity/inattention and peer problems compared to the no FI group. Low FI – reflecting marginal food security – also associated with heightened peer problems, suggesting a dose-response relationship. Greatest differences between food-secure and food-insecure children in this study occurred in late childhood and mid-adolescence. Findings additionally highlight that the impact of FI has remained consistent for UK children born in the early 1990s and mid-2000s, despite secular changes in youth mental health problems.

*Implications of all the available evidence:* Childhood food insecurity casts a long shadow on mental health and well-being outcomes throughout the lifespan, as demonstrated by substantial work from the 20^th^ and 21^st^ centuries. Here, we also see that the impact of FI on children and young people’s mental health has remained consistent over two generations of UK youth. If left unchecked, the growing prevalence of FI in the UK will only intensify concerns for the long-term mental health of the nation’s young people – especially those from low-income and marginalised backgrounds who already face a widening gap in health inequalities.

## Introduction

Food insecurity (FI) – the lack of reliable access to sufficient safe, nutritious food – has widespread negative impacts on child health and developmental outcomes.^1,2^ Despite this, FI remains prevalent in high-income countries such as the United Kingdom (UK).^3,4^ The UK has seen a sharp increase in food bank usage over the past decade,^4,5^ as rising costs of living have forced more households (including those who were previously financially stable) to seek emergency food aid.^6^ Young children continue to face disproportionately high rates of FI,^7,8^ furthering concerns about its potential impact on children and young people’s mental health in the longer-term.

FI encompasses a range of experiences depending on severity: anxiety about accessing adequate food (marginal food security), reduced diet quality, variety and desirability (low food security) and in severe cases, reduced food intake and hunger (very low food security).^9^ While FI exists on a continuum, existing literature has largely utilised a binary categorisation, leaving the potential psychological impacts of marginal food security relatively under-investigated. Emerging evidence, however, suggests a dose-response effect where odds (and severity) of mental health problems, depression and suicidality increase linearly with FI severity.^1,10,11^ Finally, while closely tied to poverty, previous work has demonstrated that associations between FI and mental health outcomes persist independent of confounders such as income, parental mental health and socioeconomic status.^1,2,12,13^

As the scale of FI has increased in the UK, the prevalence of mental health problems in children and young people has also risen sharply.^14,15^ Common mental health problems are a leading cause of disability worldwide,^16^ and have serious impacts on young people’s social development, educational attainment, mental health and quality of life in adulthood.^15,17,18^ However, symptoms also fluctuate across development, and clinical courses show high heterogeneity. Investigating population-averaged trajectories using repeated measures of mental health symptoms can therefore provide insight into the timing, development and duration of mental health problems over sensitive developmental periods such as adolescence.^19–21^

Though substantial work has demonstrated an association between FI and overall mental health and well-being in childhood and adolescence,^1,2,13^ less work has examined how and when these outcomes emerge and change in young people. While FI (as a binary variable) has been associated with persistently high depression symptoms over one year in South African youth aged 16-24,^22^ and persistently high hyperactivity/inattention problems between ages 4-8 years in Canada,^23^ null findings have also been reported when examining changes in FI status and internalising/externalising problems in US children.^24^ As such, relatively little is known about how childhood FI severity may shape the onset, timing, and persistence of mental health problems beyond childhood and over longer periods of follow-up.

Moreover, it is unclear whether these associations have remained consistent over time and are independent of generational differences, as mental health inequalities have only widened over recent years.^14,15,25^ Addressing the above and identifying points of divergence between mental health trajectories – e.g. where food-insecure children are most different from their peers – may additionally highlight windows of opportunity for truly effective intervention.

Here, we aimed to characterise the mental health trajectories of young people with differential exposure to FI in childhood. In contrast to the often binary classification of FI status, we examined three levels of potential FI exposure based on parent-reported difficulties in affording food at child age ∼5 years: No FI, Low FI and High FI. Using over a decade of longitudinal data from two UK birth cohorts born in the 20^th^ and 21^st^ centuries – the Avon Longitudinal Study of Parents and Children (ALSPAC; born 1990-1992) and Growing Up in Scotland (GUS; born 2004-2005) – we modelled trajectories of conduct problems, emotional symptoms, hyperactivity/inattention, and peer problems between ages 5-18 years. We then tested group differences in mental health scores for each FI level at ages 5 (GUS only), 7, 9, 11, 13 and 15 years to identify points of greatest divergence in each cohort. Models were adjusted for child sex (assigned at birth), in addition to household income, maternal mental health and neighbourhood deprivation due to the close coupling of FI with socioeconomic status and parental mental health.

Consistent with existing work, we hypothesised that High FI would be associated with higher (worse) overall trajectories of mental health problems. In contrast, we had no prior hypotheses on whether trajectories would differ between the Low FI and High/No FI groups, if differential exposure to FI may influence trajectory shape, or if differences between No, Low and High FI groups would be consistent over childhood and adolescence. We also had no prior hypotheses about the consistency of results across cohorts, as participants were born 12-14 years apart and in different countries within the UK.

## Materials and Methods

### Avon Longitudinal Study of Parents and Children

The Avon Longitudinal Study of Parents and Children (ALSPAC) recruited pregnant women living in Avon, England with estimated delivery dates between 1^st^ April 1991 – 31^st^ December 1992.^26^ During the initial recruitment period (1990-1992), ALSPAC recruited 14,541 pregnant women through media campaigns and invitation cards distributed via antenatal and maternal health services, resulting in 13,988 children who were alive at age 1.^27^ Additional eligible children were recruited after age 7, resulting in a total sample size of 15,447 pregnancies. Further details on ALSPAC recruitment and data collection are given in the Appendix. Please note that the study website contains details of all the data that is available through a fully searchable data dictionary and variable search tool: www.bristol.ac.uk/alspac/researchers/our-data/. Ethical approval for the study was obtained from the ALSPAC Ethics and Law Committee and the Local Research Ethics Committees. We estimated ALSPAC trajectories over 5 timepoints (measured between 1998-2009), from age ∼7 (KQ, 1998/99; n = 8,515) to age ∼17 years (TC, 2008/09; n = 5,720).

### Growing Up in Scotland

The present study utilised data from Growing up in Scotland (GUS) Birth Cohort 1, a nationally representative sample of children born in Scotland between 1^st^ June 2004 and 31^st^ May 2005.^28^ Families with eligible children were randomly sampled from Child Benefit records to receive an invitation letter; 5,217 children were enrolled when data collection began in 2005. An additional 502 children were recruited at age 9 to improve sample representativeness; further details on recruitment and data collection can be found on the study website: https://growingupinscotland.org.uk/. Ethical approval for GUS sweeps 1-8 was granted by the Scotland “A” MREC committee (reference: 04/M RE 1 0/59); for sweeps 9, 10 and beyond, ethical approval was granted by the NatCen Research Ethics Committee. We used 6 timepoints from GUS, from age ∼5 (sweep 5, 2009/10; n = 3,833) to age ∼15 years (sweep 10, 2019/2020; n = 2,669).

### Choice of primary measure

Mental health outcomes were parent-reported in both cohorts using the Strengths and Difficulties Questionnaire (SDQ), a brief screening tool for psychosocial problems in 3- 16 year olds. We estimated trajectories for four SDQ subscales: Conduct Problems, Emotional Symptoms, Hyperactivity/Inattention and Peer Problems.^29^ The SDQ was selected due to its availability in both ALSPAC and GUS, and for its use in both clinical and research settings; it is also freely available online (www.sdqinfo.org). It has demonstrated acceptable psychometric properties across cultures, correlating with other parent- and clinician-reported measures of psychopathology.^30^

### Food insecurity at age 5

Affording food is a core element across existing measures of FI.^9^ Here, we defined FI status as the degree to which parents were impacted by the cost of food, with three levels of potential severity: No FI (no impact), Low FI (slight impact, may reflect anxiety about affording food) and High FI (large impact, may reflect reduced diet quality). At child age ∼5 years, ALSPAC mothers were asked “How difficult at the moment do you find it to afford food?”; in GUS, they were asked “And how much does the cost of food affect what you give *[child name]* to eat?”. Responses were recorded on a 4-point Likert scale; we merged the two highest levels of difficulty/severity to form the “High FI” group.

### Covariates

Alongside unadjusted trajectories (i.e. age x FI status only), we fit fully-adjusted trajectories for each SDQ subscale. Covariates were collected concurrently with FI (age ∼5 years) as far as possible; if a concurrent measure was not available, we used variables from the closest timepoint *prior* to FI measurement. As FI is closely related to socioeconomic status, we included household income and area deprivation as covariates; we also controlled for maternal mental health and child sex at birth which associate with differences in mental health trajectories.

For ALSPAC analyses, we included weekly household income measured at ∼2 years 9 months postpartum (H questionnaire). Area deprivation – Index of Multiple Deprivation (IMD) quintile – and maternal mental health – Edinburgh Postnatal Depression Scale (EPDS) score – covariates were measured concurrently with FI status at ∼5 years 1 month postpartum (K questionnaire).

All GUS covariates were measured concurrently with FI status at child age ∼5 years (sweep 5). Household income was equivalised to account for household size and composition;^28^ area deprivation and maternal (primary caregiver) mental health covariates were included as Scottish Index of Multiple Deprivation (SIMD) quintile and the Short Form 12 Health Survey (SF-12), respectively.

### Non-response weighting

To correct for imbalances in non-response rates across sweeps and restore sample representativeness, data were weighted using inverse probability weights, where response at each timepoint was modelled based on sociodemographic characteristics. GUS weights were derived centrally by the Scottish Centre for Social Research and have been described previously;^31^ ALSPAC weighting is described in full in the Appendix, page 2.

### Missing data handling

Participants missing FI or covariate data and those with fewer than 2 SDQ measurements were excluded from analyses. Descriptive statistics for ALSPAC participants included (n = 6,182) and excluded (n = 8,847) from analyses are presented in Appendix Table S1; descriptive statistics for GUS participants included (n = 3,167) and excluded (n = 666) from analyses are presented in Appendix Table S2. Missing outcome data in the trajectories were handled using maximum likelihood, as per previous research.^32^

### Statistical analyses

We used multilevel growth curve models to estimate trajectories of SDQ with by-participant random intercepts and slopes.^33^ For each subscale, we first fit age-only models with increasing-order polynomials: age (linear model), age^2^ (quadratic model), age^3^ (cubic model) and age^4^ (quartic model). The best-fitting trajectory for each SDQ subscale was determined by comparing model fit indices (AIC, BIC and deviance) and likelihood ratio tests comparing each model to the one prior (with a lower-order polynomial). Age-only model results are presented in Appendix Figures S1-S2 and Tables S3-4.

Next, we stratified trajectories by FI level, including interactions between FI level and age (including polynomial terms), as group trajectories may differ in the timing and velocity of changes in their overall scores. Results from unadjusted models are presented in Appendix Figures S3-4, Tables S5-S8 and S14-16. Finally, we fit fully-adjusted models with covariates of child sex assigned at birth, household income, maternal mental health and area-level deprivation. Model estimates for fully-adjusted trajectories are presented in Appendix Tables S9-12 and S17-20.

Mean SDQ scores at ages 5 (GUS only), 7, 9, 11, 13 and 15 years were predicted within each FI group, averaging over covariate levels (i.e. marginal means). We conducted pairwise comparisons of (1) Low vs. No FI, (2) High vs. No FI, and (3) High vs Low FI scores using linear contrasts; *p*-values were adjusted for multiple testing using False Discovery Rate correction and are presented as *p*_adj_ in-text. Additional technical details of parameters used to estimate trajectories and marginal means are presented in the Appendix, page 3.

All statistical analyses were conducted in R version 4.5.0;^34^ R code used for analyses are openly available on GitHub (https://github.com/EileenYXu/FI_Trajectories).

## Results

Demographic information for ALSPAC and GUS samples are available in Table 1. Both cohorts had slightly more male than female participants (ALSPAC 50.97%; GUS 51.56% Male) and were predominately of white ethnicity (ALSPAC 96.32%; GUS 97.06%). While the majority of households in both cohorts owned their home (ALSPAC 83.92%; GUS 74.85%), fewer lived in subsidised or social housing in ALSPAC (10.57%) compared to GUS (20.06%). Similarly, fewer ALSPAC households lived in the most deprived areas (9.06% in most deprived quintile) compared to GUS (16.45%); more ALSPAC households resided in the least deprived areas (33.1% in least deprived quintile) than in GUS (23.49%).

**Table 1:** Sample demographics.

|  |  | **ALSPAC (n = 6182)** | **GUS (n = 3167)** |
| --- | --- | --- | --- |
| Child sex at birth | Male | 3151 (50.97%) | 1633 (51.56%) |
|  | Female | 3031 (49.03%) | 1534 (48.44%) |
| Income <sup>a</sup> | Lowest | 427 (6.91%) | 583 (18.41%) |
|  | 2 | 1022 (16.53%) | 698 (22.04%) |
|  | 3 | 1770 (28.63%) | 585 (18.47%) |
|  | 4 | 1403 (22.69%) | 768 (24.25%) |
|  | Highest | 1560 (25.23%) | 533 (16.83%) |
| FI level | No FI | 4972 (80.43%) | 1877 (59.27%) |
|  | High FI | 329 (5.32%) | 334 (10.55%) |
|  | Low FI | 881 (14.25%) | 956 (30.19%) |
| Area deprivation | Least deprived | 2046 (33.1%) | 744 (23.49%) |
|  | 2 | 1548 (25.04%) | 714 (22.54%) |
|  | 3 | 1071 (17.32%) | 645 (20.37%) |
|  | 4 | 957 (15.48%) | 543 (17.15%) |
|  | Most deprived | 560 (9.06%) | 521 (16.45%) |
| Maternal education <sup>b</sup> | >=A level | 2568 (42.33%) | 2147 (67.84%) |
|  | <O level/GCSE | 1313 (21.65%) | 194 (6.13%) |
|  | O level/GCSE | 2185 (36.02%) | 824 (26.03%) |
| Occupational class <sup>c</sup> | I-II | 2156 (41.2%) | 1358 (42.89%) |
|  | III-V | 3077 (58.8%) | 1808 (57.11%) |
| Parity <sup>d</sup> | 1st born | 2816 (46.63%) | 1529 (48.28%) |
|  | 2nd born | 2189 (36.25%) | 1079 (34.07%) |
|  | 3rd + | 1034 (17.12%) | 559 (17.65%) |
| Housing tenure | Mortgage/owned | 4947 (83.92%) | 2306 (74.85%) |
|  | Private rented | 325 (5.51%) | 157 (5.1%) |
|  | Subsidised rented | 623 (10.57%) | 618 (20.06%) |
| Child ethnicity <sup>e</sup> | White | 5757 (96.32%) | 3072 (97.06%) |
|  | Non-white | 220 (3.68%) | 93 (2.94%) |
| Smoking during pregnancy <sup>f</sup> | No | 4853 (79.52%) | 2492 (79.92%) |
|  | Yes | 1250 (20.48%) | 626 (20.08%) |

Sample demographics of ALSPAC and GUS participants included in analyses.
ALSPAC = Avon Longitudinal Study of Parents and Children; FI = food insecurity; GUS = Growing Up in Scotland.
<sup>a</sup> In ALSPAC, weekly income bands were: <£100 (Lowest); £100 to £199 (2); £200 to £299 (3); £300 to £399 (4); >£400 (Highest). GUS used annual income equivalised for household composition: <£12,217 (Lowest); >=£12,217 to <£19,643 (2); >=£19,643 to <£29,126 (3); >=£29,126 to <£37,857 (4); >=£37,857 (Highest).
<sup>b</sup> O level in ALSPAC, GCSE in GUS.
<sup>c</sup> ALSPAC defined occupational class using the UK Registrar General's occupational coding (SOC 90). In GUS, occupational class was defined using the National Statistics Socio-economic Classification (NS-SEC); classes I-II correspond to managerial, administrative and professional occupations; classes III-V correspond to intermediate occupations to unemployed.
<sup>d</sup> Birth order in GUS.
<sup>e</sup> In ALSPAC, child ethnicity was derived from mother reports of her and her partner's ethnic backgrounds. Child ethnicity was coded as non-white if either parent had a non-white background, which included: Black Caribbean, Black African, Other black, Indian, Pakistani, Bangladeshi, Chinese, Other.
In GUS, non-white ethnicity includes the following: Mixed - White and Black Caribbean, Mixed - White and Black African, Mixed - White and Asian, Any other mixed background, Asian/Asian British – Indian, Asian/Asian British – Pakistani, Asian/Asian British – Bangladeshi, Any other Asian/Asian British background, Black/Black British – Caribbean, Black/Black British – African, Any other Black/Black British background, Chinese, Any other non-white background.
<sup>f</sup> ALSPAC measured smoking during the first trimester; GUS measured smoking over the entire pregnancy.

At child age 5, the majority (80.43%) of ALSPAC mothers reported no difficulty in affording food (No FI). 14.25% reported slight difficulty (Low FI), and 5.32% found it fairly or very difficult (High FI). In GUS, fewer respondents were unaffected by the cost of food (59.27% No FI). 30.19% reported that the cost of food had “a little” impact on what they gave their child to eat (Low FI), with 10.55% of respondents being impacted “a fair amount” or “a lot” (High FI).

After controlling for covariates, High FI groups demonstrated higher trajectories for all SDQ subscales and across both cohorts (Figure 1). Mean SDQ scores derived from the trajectories for each subscale, averaged over all covariate levels, are displayed by age and FI level in Table 2; Tables 3 and 4 show pairwise comparisons between scores for each FI level in ALSPAC and GUS, respectively. Results for individual symptom trajectories are described below; findings which were replicated across both cohorts are prioritised in the main text for brevity. Additional findings from ALSPAC and GUS analyses are described in the Appendix, pages 3-6; for completeness, model coefficients are presented in Appendix Tables S5-S20.

**Figure 1:**
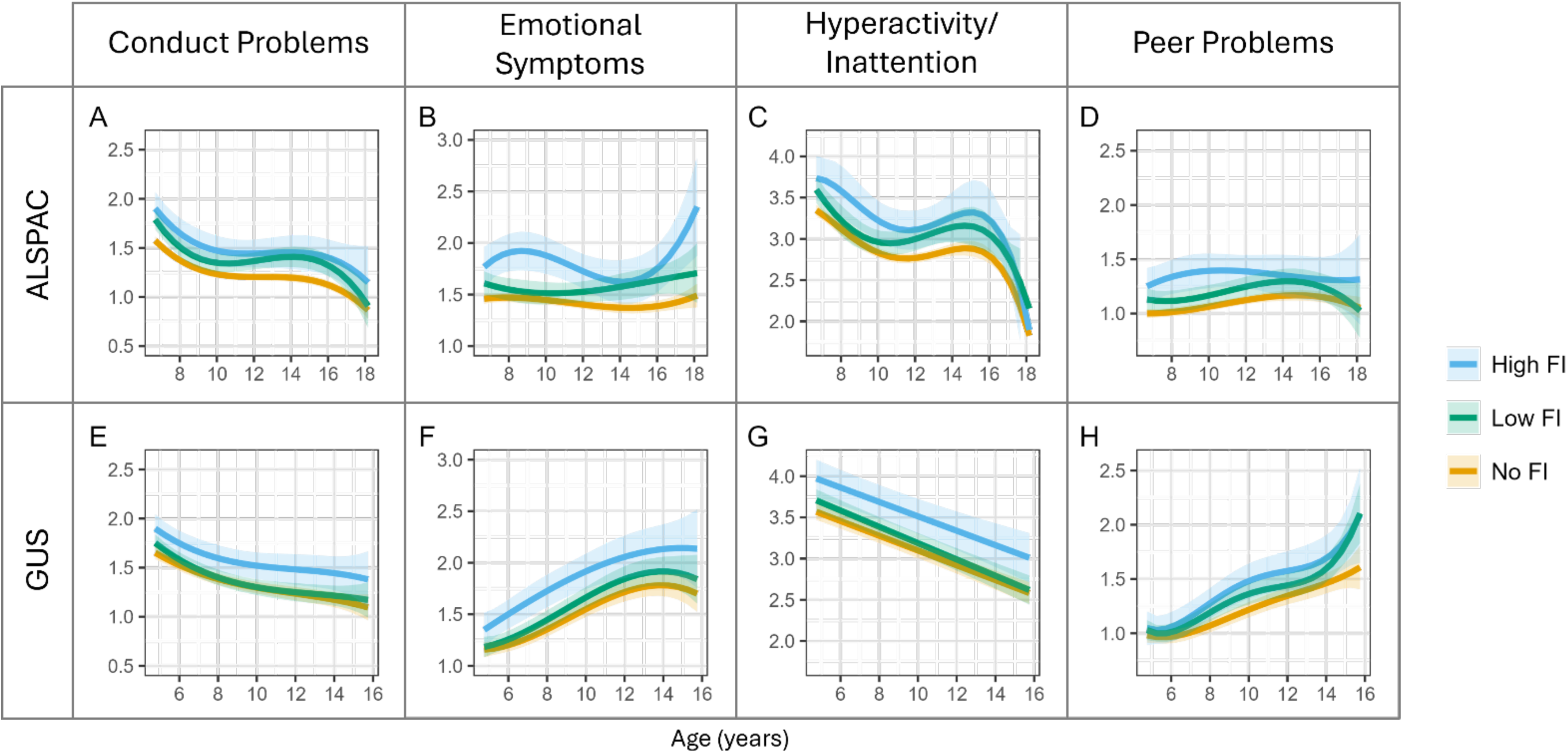
Fully-adjusted SDQ trajectories by FI status. Mean trajectories of SDQ subscale scores in ALSPAC (A-D) and GUS (E-H), stratified by food insecurity exposure at age 5. Plotted trajectories for each group are averaged over covariate levels. ALSPAC = Avon Longitudinal Study of Parents and Children; FI = food insecurity; GUS = Growing Up in Scotland; SDQ = Strengths and Difficulties Questionnaire.

**Table 2:** SDQ subscale scores by FI status.

| Age | FI Level | Conduct Problems |  | Emotional Problems |  | Hyperactivity |  | Peer Problems |  |
| --- | --- | --- | --- | --- | --- | --- | --- | --- | --- |
|  |  | ALSPAC | GUS | ALSPAC | GUS | ALSPAC | GUS | ALSPAC | GUS |
| 5 | No FI |  | 1.62 [1.56 - 1.68] |  | 1.16 [1.10 - 1.23] |  | 3.55 [3.45 - 3.64] |  | 0.97 [0.91 - 1.03] |
|  | Low FI |  | 1.71 [1.63 - 1.79] |  | 1.19 [1.10 - 1.28] |  | 3.68 [3.55 - 3.81] |  | 1.01 [0.93 - 1.09] |
|  | High FI |  | 1.86 [1.73 - 2.00] |  | 1.38 [1.23 - 1.53] |  | 3.95 [3.72 - 4.17] |  | 1.03 [0.90 - 1.17] |
| 7 | No FI | 1.51 [1.48 - 1.55] | 1.45 [1.39 - 1.50] | 1.46 [1.42 - 1.51] | 1.27 [1.20 - 1.33] | 3.30 [3.23 - 3.36] | 3.37 [3.28 - 3.46] | 1.00 [0.96 - 1.04] | 1.01 [0.95 - 1.07] |
|  | Low FI | 1.70 [1.61 - 1.80] | 1.48 [1.40 - 1.56] | 1.59 [1.48 - 1.70] | 1.34 [1.25 - 1.43] | 3.49 [3.33 - 3.65] | 3.48 [3.36 - 3.61] | 1.12 [1.03 - 1.21] | 1.09 [1.00 - 1.17] |
|  | High FI | 1.83 [1.67 - 1.98] | 1.66 [1.53 - 1.79] | 1.82 [1.64 - 2.00] | 1.62 [1.47 - 1.77] | 3.72 [3.46 - 3.98] | 3.77 [3.57 - 3.98] | 1.28 [1.12 - 1.43] | 1.15 [1.01 - 1.29] |
| 9 | No FI | 1.28 [1.24 - 1.32] | 1.33 [1.28 - 1.39] | 1.46 [1.42 - 1.51] | 1.45 [1.38 - 1.52] | 2.95 [2.89 - 3.02] | 3.19 [3.10 - 3.28] | 1.03 [0.99 - 1.08] | 1.14 [1.08 - 1.21] |
|  | Low FI | 1.40 [1.30 - 1.49] | 1.34 [1.26 - 1.42] | 1.52 [1.41 - 1.64] | 1.55 [1.46 - 1.65] | 3.05 [2.88 - 3.22] | 3.29 [3.17 - 3.41] | 1.13 [1.03 - 1.23] | 1.29 [1.20 - 1.38] |
|  | High FI | 1.53 [1.38 - 1.69] | 1.55 [1.42 - 1.68] | 1.92 [1.73 - 2.11] | 1.83 [1.67 - 1.99] | 3.39 [3.12 - 3.67] | 3.60 [3.39 - 3.81] | 1.38 [1.21 - 1.54] | 1.39 [1.24 - 1.54] |
| 11 | No FI | 1.21 [1.17 - 1.24] | 1.26 [1.20 - 1.33] | 1.43 [1.38 - 1.47] | 1.64 [1.56 - 1.72] | 2.77 [2.71 - 2.83] | 3.01 [2.91 - 3.10] | 1.09 [1.06 - 1.13] | 1.28 [1.21 - 1.36] |
|  | Low FI | 1.34 [1.26 - 1.43] | 1.27 [1.18 - 1.36] | 1.51 [1.41 - 1.61] | 1.76 [1.65 - 1.87] | 2.95 [2.81 - 3.09] | 3.09 [2.96 - 3.22] | 1.21 [1.12 - 1.30] | 1.41 [1.31 - 1.51] |
|  | High FI | 1.45 [1.30 - 1.59] | 1.50 [1.35 - 1.64] | 1.81 [1.64 - 1.97] | 1.99 [1.80 - 2.18] | 3.12 [2.89 - 3.36] | 3.42 [3.20 - 3.65] | 1.40 [1.25 - 1.55] | 1.54 [1.37 - 1.71] |
| 13 | No FI | 1.20 [1.17 - 1.24] | 1.21 [1.13 - 1.28] | 1.38 [1.34 - 1.43] | 1.77 [1.67 - 1.86] | 2.81 [2.75 - 2.87] | 2.83 [2.72 - 2.94] | 1.15 [1.11 - 1.19] | 1.40 [1.31 - 1.49] |
|  | Low FI | 1.39 [1.30 - 1.49] | 1.23 [1.14 - 1.33] | 1.55 [1.43 - 1.66] | 1.90 [1.77 - 2.03] | 3.07 [2.93 - 3.22] | 2.89 [2.75 - 3.04] | 1.28 [1.18 - 1.38] | 1.49 [1.37 - 1.61] |
|  | High FI | 1.45 [1.30 - 1.61] | 1.47 [1.30 - 1.63] | 1.66 [1.47 - 1.84] | 2.10 [1.88 - 2.33] | 3.17 [2.93 - 3.41] | 3.25 [3.00 - 3.50] | 1.37 [1.20 - 1.54] | 1.61 [1.41 - 1.81] |
| 15 | No FI | 1.18 [1.13 - 1.22] | 1.13 [1.03 - 1.23] | 1.37 [1.32 - 1.43] | 1.75 [1.62 - 1.89] | 2.88 [2.79 - 2.98] | 2.65 [2.53 - 2.77] | 1.17 [1.12 - 1.22] | 1.53 [1.42 - 1.65] |
|  | Low FI | 1.39 [1.28 - 1.50] | 1.19 [1.06 - 1.32] | 1.61 [1.47 - 1.74] | 1.89 [1.71 - 2.07] | 3.16 [2.92 - 3.39] | 2.69 [2.53 - 2.86] | 1.29 [1.17 - 1.41] | 1.82 [1.66 - 1.97] |
|  | High FI | 1.44 [1.26 - 1.62] | 1.41 [1.20 - 1.63] | 1.65 [1.42 - 1.87] | 2.14 [1.85 - 2.43] | 3.32 [2.94 - 3.71] | 3.08 [2.79 - 3.36] | 1.33 [1.13 - 1.53] | 1.86 [1.60 - 2.12] |

Mean [95% CI] predicted SDQ subscale scores at age 5 (GUS only), 7, 9, 11, 13 and 15 years, averaging over all covariate levels.
ALSPAC = Avon Longitudinal Study of Parents and Children; FI = food insecurity; GUS = Growing Up in Scotland; SDQ = Strengths and Difficulties Questionnaire.

**Table 3:**
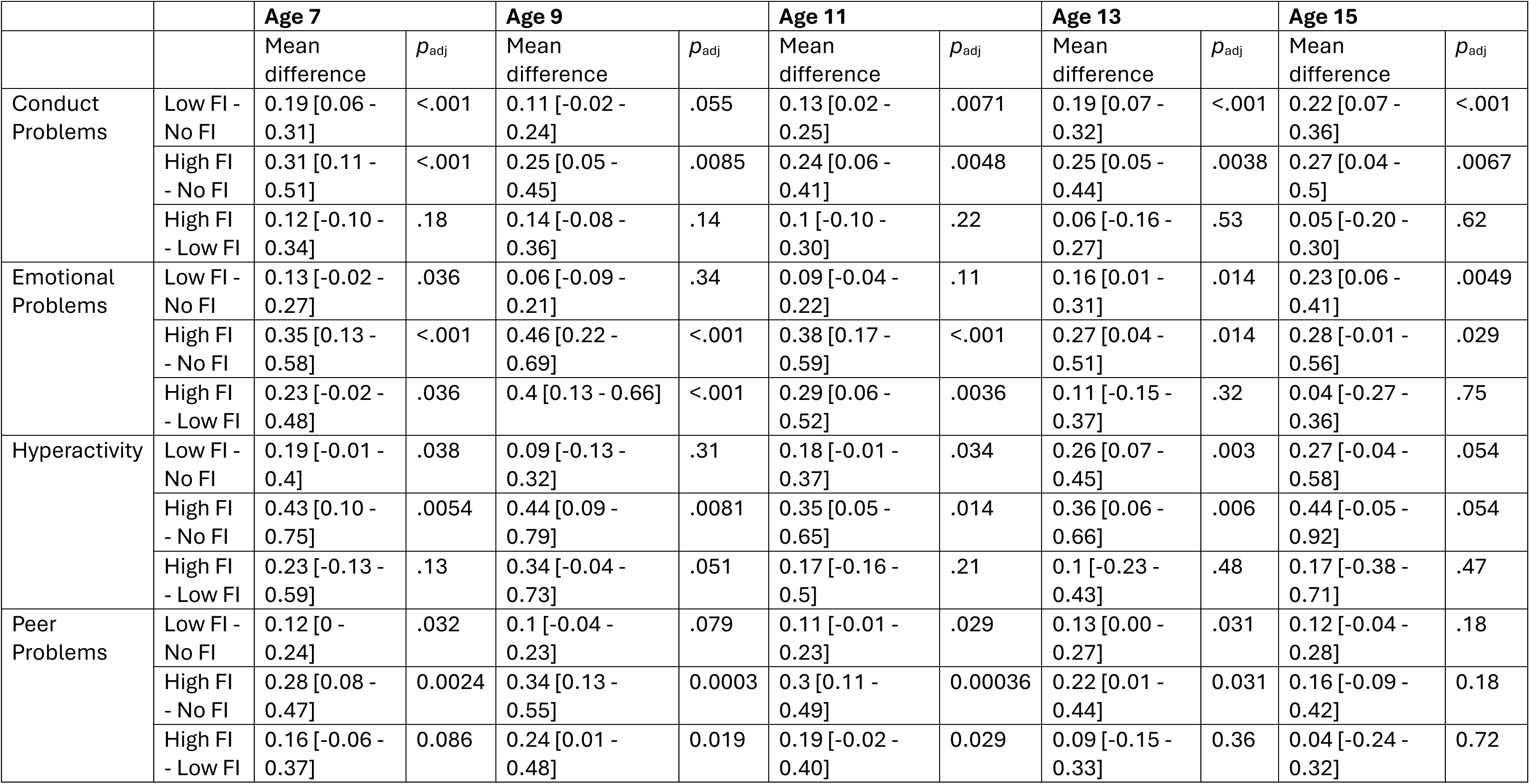
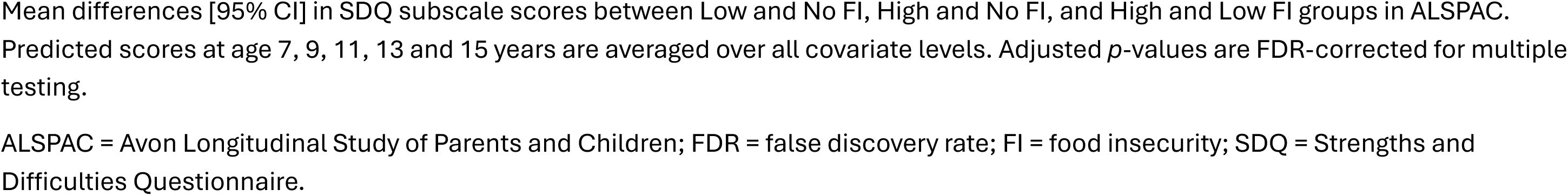
Pairwise comparisons of SDQ scores in ALSPAC.

|  |  | **Age 7** |  | **Age 9** |  | **Age 11** |  | **Age 13** |  | **Age 15** |  |
| --- | --- | --- | --- | --- | --- | --- | --- | --- | --- | --- | --- |
| | | Mean difference | $p_{adj}$ | Mean difference | $p_{adj}$ | Mean difference | $p_{adj}$ | Mean difference | $p_{adj}$ | Mean difference | $p_{adj}$ |
| Conduct Problems | Low FI - No FI | 0.19 [0.06 - 0.31] | <.001 | 0.11 [-0.02 - 0.24] | .055 | 0.13 [0.02 - 0.25] | .0071 | 0.19 [0.07 - 0.32] | <.001 | 0.22 [0.07 - 0.36] | <.001 |
|  | High FI - No FI | 0.31 [0.11 - 0.51] | <.001 | 0.25 [0.05 - 0.45] | .0085 | 0.24 [0.06 - 0.41] | .0048 | 0.25 [0.05 - 0.44] | .0038 | 0.27 [0.04 - 0.5] | .0067 |
|  | High FI - Low FI | 0.12 [-0.10 - 0.34] | .18 | 0.14 [-0.08 - 0.36] | .14 | 0.1 [-0.10 - 0.30] | .22 | 0.06 [-0.16 - 0.27] | .53 | 0.05 [-0.20 - 0.30] | .62 |
| Emotional Problems | Low FI - No FI | 0.13 [-0.02 - 0.27] | .036 | 0.06 [-0.09 - 0.21] | .34 | 0.09 [-0.04 - 0.22] | .11 | 0.16 [0.01 - 0.31] | .014 | 0.23 [0.06 - 0.41] | .0049 |
|  | High FI - No FI | 0.35 [0.13 - 0.58] | <.001 | 0.46 [0.22 - 0.69] | <.001 | 0.38 [0.17 - 0.59] | <.001 | 0.27 [0.04 - 0.51] | .014 | 0.28 [-0.01 - 0.56] | .029 |
|  | High FI - Low FI | 0.23 [-0.02 - 0.48] | .036 | 0.4 [0.13 - 0.66] | <.001 | 0.29 [0.06 - 0.52] | .0036 | 0.11 [-0.15 - 0.37] | .32 | 0.04 [-0.27 - 0.36] | .75 |
| Hyperactivity | Low FI - No FI | 0.19 [-0.01 - 0.4] | .038 | 0.09 [-0.13 - 0.32] | .31 | 0.18 [-0.01 - 0.37] | .034 | 0.26 [0.07 - 0.45] | .003 | 0.27 [-0.04 - 0.58] | .054 |
|  | High FI - No FI | 0.43 [0.10 - 0.75] | .0054 | 0.44 [0.09 - 0.79] | .0081 | 0.35 [0.05 - 0.65] | .014 | 0.36 [0.06 - 0.66] | .006 | 0.44 [-0.05 - 0.92] | .054 |
|  | High FI - Low FI | 0.23 [-0.13 - 0.59] | .13 | 0.34 [-0.04 - 0.73] | .051 | 0.17 [-0.16 - 0.5] | .21 | 0.1 [-0.23 - 0.43] | .48 | 0.17 [-0.38 - 0.71] | .47 |
| Peer Problems | Low FI - No FI | 0.12 [0 - 0.24] | .032 | 0.1 [-0.04 - 0.23] | .079 | 0.11 [-0.01 - 0.23] | .029 | 0.13 [0.00 - 0.27] | .031 | 0.12 [-0.04 - 0.28] | .18 |
|  | High FI - No FI | 0.28 [0.08 - 0.47] | 0.0024 | 0.34 [0.13 - 0.55] | 0.0003 | 0.3 [0.11 - 0.49] | 0.00036 | 0.22 [0.01 - 0.44] | 0.031 | 0.16 [-0.09 - 0.42] | 0.18 |
|  | High FI - Low FI | 0.16 [-0.06 - 0.37] | 0.086 | 0.24 [0.01 - 0.48] | 0.019 | 0.19 [-0.02 - 0.40] | 0.029 | 0.09 [-0.15 - 0.33] | 0.36 | 0.04 [-0.24 - 0.32] | 0.72 |

Mean differences [95% CI] in SDQ subscale scores between Low and No FI, High and No FI, and High and Low FI groups in ALSPAC. Predicted scores at age 7, 9, 11, 13 and 15 years are averaged over all covariate levels. Adjusted $p$ -values are FDR-corrected for multiple testing.
ALSPAC = Avon Longitudinal Study of Parents and Children; FDR = false discovery rate; FI = food insecurity; SDQ = Strengths and Difficulties Questionnaire.

**Table 4:** Pairwise comparisons of SDQ scores in GUS.

|  |  | **Age 5** |  | **Age 7** |  | **Age 9** |  | **Age 11** |  | **Age 13** |  | **Age 15** |  |
| --- | --- | --- | --- | --- | --- | --- | --- | --- | --- | --- | --- | --- | --- |
| | | Mean difference | $p_{adj}$ | Mean difference | $p_{adj}$ | Mean difference | $p_{adj}$ | Mean difference | $p_{adj}$ | Mean difference | $p_{adj}$ | Mean difference | $p_{adj}$ |
| Conduct Problems | Low FI - No FI | 0.09 [-0.03 - 0.21] | 0.083 | 0.03 [-0.09 - 0.15] | 0.5 | 0.01 [-0.11 - 0.13] | 0.87 | 0.01 [-0.12 - 0.14] | 0.89 | 0.03 [-0.12 - 0.17] | 0.66 | 0.06 [-0.14 - 0.26] | 0.46 |
|  | High FI - No FI | 0.24 [0.05 - 0.43] | 0.0059 | 0.21 [0.04 - 0.39] | 0.012 | 0.22 [0.04 - 0.39] | 0.011 | 0.23 [0.04 - 0.43] | 0.012 | 0.26 [0.04 - 0.48] | 0.014 | 0.28 [-0.01 - 0.57] | 0.059 |
|  | High FI - Low FI | 0.15 [-0.04 - 0.35] | 0.083 | 0.18 [-0.01 - 0.37] | 0.031 | 0.21 [0.02 - 0.39] | 0.011 | 0.23 [0.02 - 0.43] | 0.012 | 0.23 [0.00 - 0.46] | 0.024 | 0.22 [-0.09 - 0.52] | 0.13 |
| Emotional Problems | Low FI - No FI | 0.03 [-0.10 - 0.17] | 0.56 | 0.08 [-0.06 - 0.21] | 0.19 | 0.1 [-0.04 - 0.25] | 0.085 | 0.12 [-0.05 - 0.29] | 0.089 | 0.13 [-0.07 - 0.33] | 0.12 | 0.14 [-0.13 - 0.41] | 0.22 |
|  | High FI - No FI | 0.22 [0.01 - 0.42] | 0.031 | 0.35 [0.15 - 0.56] | 0.00013 | 0.38 [0.16 - 0.59] | 0.000098 | 0.35 [0.1 - 0.61] | 0.0027 | 0.34 [0.04 - 0.64] | 0.02 | 0.39 [0.00 - 0.78] | 0.054 |
|  | High FI - Low FI | 0.19 [-0.03 - 0.4] | 0.057 | 0.27 [0.06 - 0.49] | 0.0034 | 0.27 [0.04 - 0.5] | 0.0064 | 0.23 [-0.04 - 0.5] | 0.058 | 0.21 [-0.11 - 0.52] | 0.12 | 0.25 [-0.17 - 0.67] | 0.22 |
| Hyperactivity | Low FI - No FI | 0.14 [-0.06 - 0.34] | 0.1 | 0.12 [-0.07 - 0.3] | 0.13 | 0.1 [-0.08 - 0.28] | 0.19 | 0.08 [-0.12 - 0.28] | 0.32 | 0.06 [-0.16 - 0.28] | 0.5 | 0.04 [-0.21 - 0.3] | 0.68 |
|  | High FI - No FI | 0.4 [0.10 - 0.71] | 0.0042 | 0.41 [0.13 - 0.69] | 0.0015 | 0.41 [0.13 - 0.69] | 0.0012 | 0.42 [0.12 - 0.72] | 0.0025 | 0.42 [0.09 - 0.76] | 0.0077 | 0.43 [0.04 - 0.81] | 0.023 |
|  | High FI - Low FI | 0.27 [-0.05 - 0.59] | 0.065 | 0.29 [0.00 - 0.58] | 0.027 | 0.31 [0.02 - 0.61] | 0.015 | 0.34 [0.02 - 0.65] | 0.015 | 0.36 [0.01 - 0.71] | 0.021 | 0.38 [-0.02 - 0.79] | 0.035 |
| Peer Problems | Low FI - No FI | 0.04 [-0.08 - 0.16] | 0.67 | 0.08 [-0.05 - 0.21] | 0.2 | 0.15 [0.01 - 0.28] | 0.016 | 0.12 [-0.03 - 0.28] | 0.073 | 0.09 [-0.1 - 0.27] | 0.3 | 0.28 [0.04 - 0.52] | 0.016 |
|  | High FI - No FI | 0.06 [-0.12 - 0.25] | 0.67 | 0.15 [-0.04 - 0.34] | 0.19 | 0.25 [0.05 - 0.45] | 0.01 | 0.25 [0.03 - 0.48] | 0.021 | 0.21 [-0.06 - 0.48] | 0.19 | 0.33 [-0.02 - 0.68] | 0.039 |
|  | High FI - Low FI | 0.02 [-0.17 - 0.22] | 0.76 | 0.07 [-0.13 - 0.27] | 0.42 | 0.1 [-0.11 - 0.32] | 0.25 | 0.13 [-0.11 - 0.37] | 0.19 | 0.12 [-0.16 - 0.41] | 0.3 | 0.04 [-0.33 - 0.42] | 0.77 |

Mean differences in SDQ subscale scores between Low and No FI, High and No FI, and High and Low FI groups in GUS. Predicted scores at age 5, 7, 9, 11, 13 and 15 years are averaged over all covariate levels. Adjusted *p*-values are FDR-corrected for multiple testing.
FDR = false discovery rate; FI = food insecurity; GUS = Growing Up in Scotland; SDQ = Strengths and Difficulties Questionnaire.

### Conduct Problems

In both cohorts, overall levels of conduct problems decreased between ages 5 and 15 (Figure 1, Appendix p 3-6). Individuals with High FI had higher conduct problem trajectories in both cohorts (Figure 1 panels A and E), with comparable increases in model intercept at the mean age across assessments (11.19 years in ALSPAC; 8.78 years in GUS) compared to No FI (β_ALSPAC_=0.24 [0.09,0.38], *p*=.001; β_GUS_=0.21 [0.07,0.36], *p*=.004).

The greatest difference in mean conduct problem scores between High vs. No FI groups occurred at age 7 (0.31 [0.11,0.51], *p*_adj_<.001) in ALSPAC. High and Low FI groups did not significantly differ at any age (Table 3); the greatest difference for Low vs. No FI occurred at age 15 (0.22 [0.07,0.36], *p*_adj_<.001). In GUS, differences in conduct problems were most pronounced at age 13 for High vs. No FI (0.26 [0.04,0.48], *p*_adj_=.014) and Low FI (0.23 [0.00,0.46], *p*_adj_=.024); Low and No FI groups showed no differences in conduct problem scores (Table 4).

### Emotional Symptoms

High FI was associated with elevated emotional symptoms compared to No FI at model intercept in both cohorts (β_ALSPAC_=0.37 [0.20,0.54] at 11.19 years, *p*<.001; β_GUS_=0.38 [0.20,0.55] at 8.78 years, *p*<.001). However, trajectories differed in overall shape (Figure 1, Appendix p 3-4). In ALSPAC, emotional symptoms were relatively stable in Low and No FI groups compared to the High FI group which showed a sharp increase in symptoms beginning in mid-adolescence (Figure 1B, Table 2). In contrast, all FI groups in GUS showed a steady increase in symptoms between ages 5-15 (Figure 1F, Table 2).

Group differences in emotional symptom scores were most pronounced at age 9 in ALSPAC for High vs. No FI (0.46 [0.22,0.69], *p*_adj_<.001) and Low FI 0.40 [0.13,0.66], *p*_adj_<.001); for Low vs. No FI this occurred at age 15 (0.23 [0.06,0.41], *p*_adj_=.005). In GUS, greatest differences also occurred at age 9 for High vs. No (0.38 [0.16,0.59], *p*_adj_<.001) and Low FI groups (0.27 [0.04,0.5], *p*_adj_=.006). Low and No FI groups did not differ at any age in GUS (Table 4).

### Hyperactivity/Inattention

Levels of hyperactivity/inattention decreased over childhood and adolescence in both cohorts (Figure 1, Appendix p 3-6), though ALSPAC trajectories showed a small peak in scores during mid-adolescence (Figure 1C, Appendix p 3, 11). Individuals in the High FI group had elevated trajectories compared to No FI in both cohorts; this increase in model intercept was less pronounced in ALSPAC (β=0.35 [0.10,0.59] at 11.19 years, *p*=.005) than in GUS (β=0.41 [0.18,0.64] at 8.78 years, *p*<.001).

Group differences in hyperactivity/inattention scores were most pronounced at age 9 for High vs. No FI in ALSPAC (0.44 [0.09,0.79], *p*_adj_=.008) and at age 13 for Low vs. No FI groups (0.26 [0.07,0.45], *p*_adj_=.003); High and Low FI groups did not differ at any age (Table 3). In GUS, differences were greatest at age 15 for High vs. No FI (0.43 [0.04,0.81], *p*_adj_=.023) and age 13 for High vs. Low FI (0.36 [0.01,0.71], *p*_adj_=.021); Low and No FI scores did not differ (Table 4).

### Peer Problems

Both High and Low FI were associated with greater levels of peer problems in both cohorts, with High FI showing a more pronounced increase in model intercept (β_ALSPAC_=0.30 [0.14,0.45] at 11.19 years, *p*<.001; β_GUS_=0.24 [0.08,0.41] 8.78 years, *p*=.004) than Low FI at the same age (β_ALSPAC_=0.12 [0.02,0.21], *p*=.021; β_GUS_=0.14 [0.03,0.25], *p*=.011). Peer problem scores also followed different trajectories in each cohort (Appendix p3-4, p11-12), remaining relatively stable over all FI levels in ALSPAC (Figure 1D). In GUS, peer problems instead rose steadily from age 5 onwards for all FI groups (Figure 1H).

In ALSPAC, differences in peer problem scores were greatest at age 9 for High vs No FI (0.34 [0.13,0.55], *p*_adj_<.001) and High vs Low FI (0.24 [0.01,0.48], *p*_adj_=.019) groups; Low and No FI groups differed most at age 13 (0.13 [0.00,0.27], *p*=.031). Mean peer problem scores did not differ between High and Low FI groups at any age in GUS (Table 4); differences were most pronounced at age 9 for High vs No FI (0.25 [0.05,0.45], *p*_adj_=.01) and at age 15 for Low vs No FI (0.28 [0.04,0.52], *p*_adj_=.025) groups.

## Discussion

In this study, we characterised 10-year trajectories of conduct, emotional, hyperactivity/inattention and peer problems across childhood and adolescence following differential exposure to childhood FI. Findings were consistent for children born in the early 1990s and mid-2000s despite generational differences. FI at age ∼5 years was associated with higher (worse) overall trajectories, even after adjusting for child sex, household income, maternal mental health problems and area deprivation. This was most pronounced for children whose parents were most heavily impacted by the cost of food (High FI), who had greater initial SDQ scores across all domains compared to their food-secure peers. This gap between trajectories remained remarkably consistent across children born in the early 1990s (ALSPAC) and mid-2000s (GUS), persisting for up to 10 years. In terms of dose-response effects, children within the Low FI group also showed higher levels of peer problems in both cohorts compared to food-secure children.

These findings corroborate previous cross-sectional and longitudinal work from the 20^th^ and 21^st^ centuries which highlight the deleterious effects of FI on child and adolescent mental health.^1,2,22,23^ By integrating repeated measurements of mental health symptoms over a 10-year period, we provide further evidence that the impact of FI is both pervasive and sustained across development – especially in High FI groups. Results from the Low FI groups are less directly comparable with existing literature, as most previous studies dichotomise FI status as either food secure or food insecure (low/very low food security).^10^ However, converging evidence suggests that marginal food security is also associated with depression, anxiety and interpersonal difficulties – consistent with findings for peer problem trajectories in our study.^1,35,36^

We found that differences in SDQ scores between food-insecure and food-secure youth were especially pronounced from age 9 onwards, towards the end of primary school education. Though parents may attempt to shield children from FI, previous work exploring children’s experiences highlight that even elementary school-aged children (5- 11 years) are often aware of – and may even take responsibility for managing food resources in the home.^37^ Alongside shouldering this stress and worry for their parents, young people also report feelings of shame and alienation from their peers.^2,37^ These may be particularly detrimental to mental health as children approach adolescence and become increasingly attuned to their social environments.

From a parental perspective, the lived experiences associated with FI and financial hardship – e.g. being forced to choose between buying food and paying for other essentials such as housing, utility bills and clothing^8,38^ – also come with substantial stress and a draining of cognitive resources needed for parenting. This, in turn, impairs parenting practices and parent-child interactions, ^38,39^ impacting child mental health in the longer-term. Given that FI is inextricably tied to social inequalities which have only widened in recent years,^7,8,25^ the detrimental impact of FI on youth mental health must be addressed in tandem with issues of income, employment and stable housing. In other words, meaningful change necessitates that all families can afford the essential costs of living and have the resources (e.g. time, skills) to buy and prepare nutritious food.^8,40^

### Strengths and Limitations

Results were successfully replicated across two large cohort studies, despite differences in sample recruitment, demographic composition, geographical location and secular trends in the prevalence of mental health problems across generations.^14,15^ This strengthens this study’s findings, which further existing FI literature by incorporating trajectories of mental health outcomes for the first time, allowing for greater inferences on onset and change over time, vital for identifying critical periods for intervention. Analyses adjusted for confounders at the time of FI exposure which was parent-reported using similar measures at the same age in each cohort; the prospective design minimises recall bias.^41^ We also harmonised our outcome measures; the SDQ in in both ALSPAC and GUS, reducing biases that may arise due to inconsistencies in measurement.

This work, however, is not without limitations. Though data were weighted to improve representativeness, reducing biases due to differential non-response,^31^ children in both cohorts were predominantly of white ethnicity. ALSPAC participants were more affluent than other parts of the UK at the time of enrolment.^26,27^ GUS achieved a more nationally representative sample, however young mothers residing in the most deprived regions of Scotland were under-represented.^28^ Thus, while these findings may be generalisable to a more privileged subset of the UK population, they are less generalisable to children from underserved groups who experience greater structural inequalities and higher rates of FI.^7,8^ Additionally, we note that the widening confidence intervals towards the end of each trajectory may reflect a reduction in statistical power as sample sizes declined. We therefore caution against extrapolating these findings beyond the sample age range, and suggest careful interpretation of trajectories beyond age 16 for this reason.

We also highlight some methodological limitations that should be addressed in future work. As this was an opportunistic study analysing previously collected data, we used a single-item measure as a proxy for FI. While affording food is a central theme across FI measures,^9^ experience-based scales of food security provide a more complete assessment of FI severity which should be prioritised in future work. However, similar to reporter discrepancies in adolescent mental health problems, parents and children have differing experiences of household FI.^37^ Children’s feelings of worry, responsibility and awareness of FI may be especially pertinent for FI, and further investigation using child-reported measures of FI and mental health symptoms is warranted. Additional avenues of investigation include integrating electronic health record data to cover longer periods of follow-up, and linking mental health and FI trajectories to future social and educational outcomes.

In conclusion, these findings provide further evidence of the detrimental impact of food insecurity on child and adolescent mental health. In two generations of UK children born in England and Scotland, FI at age 5 was consistently associated with worse conduct, emotional, hyperactivity/inattention and peer problem trajectories over the next 10 years. Across both cohorts, we also found a potential dose-response effect of FI severity on peer problem trajectories, though replication in more diverse cohorts with more robust FI measures is needed. These findings provide further evidence of the persistent negative impact of FI on child health and highlight the need for interventions to safeguard the long-term mental health of children and young people, particularly those from low-income households.

## Author contributions

EYX – Conceptualisation, Data curation, Formal analysis, Methodology, Project administration, Software, Visualisation, Writing – original draft, Writing – review & editing

AES – Data curation, Software, Writing – review & editing

ASFK – Conceptualisation, Data curation, Formal analysis, Methodology, Supervision, Writing – original draft, Writing – review & editing

HCW – Conceptualisation, Data curation, Methodology, Supervision, Writing – original draft, Writing – review & editing

## Acknowledgments

We are extremely grateful to all the families who took part in this study, the midwives for their help in recruiting them, and the whole ALSPAC team, which includes data collection staff, data and administrations staff, technical managers and the technical staff with the Bristol Bioresource Laboratory, based within the University of Bristol.

We are grateful for the entirely voluntary co-operation of the children who form the Growing Up in Scotland Cohort and their mothers, fathers and other family members, and to NatCen and to the UK Data Service for making them available. However, neither NatCen, nor the UK Data Service bear any responsibility for the analysis or interpretation of these data.

## Data availability

The informed consent obtained from ALSPAC (Avon Longitudinal Study of Parents and Children) participants does not allow the data to be made available through any third party maintained public repository. Supporting data are available from ALSPAC on request under the approved proposal number, B3421. Full instructions for applying for data access can be found here: http://www.bristol.ac.uk/alspac/researchers/access/. The ALSPAC study website contains details of all available data (http://www.bristol.ac.uk/alspac/researchers/our-data/).

GUS data can be accessed through the UK Data Service (http://doi.org/10.5255/UKDA-Series-200020). Data used in the preparation of this article are available under Special Licence Access. Further information on all available GUS data and instructions for applying for access can be found on the study website: https://growingupinscotland.org.uk/using-gus-data.

