## Supplement for "Childhood food insecurity and youth mental health trajectories in two UK longitudinal cohorts"

### Additional ALSPAC cohort information

Pregnant women resident in Avon, UK with expected dates of delivery between 1st April 1991 and 31st December 1992 were invited to take part in the study. 20,248 pregnancies have been identified as being eligible and the initial number of pregnancies enrolled was 14,541. Of the initial pregnancies, there was a total of 14,676 foetuses, resulting in 14,062 live births and 13,988 children who were alive at 1 year of age. When the oldest children were approximately 7 years of age, an attempt was made to bolster the initial sample with eligible cases who had failed to join the study originally. As a result, when considering variables collected from the age of seven onwards (and potentially abstracted from obstetric notes) there are data available for more than the 14,541 pregnancies mentioned above: The number of new pregnancies not in the initial sample (known as Phase I enrolment) that are currently represented in the released data and reflecting enrolment status at the age of 24 is 906, resulting in an additional 913 children being enrolled (456, 262 and 195 recruited during Phases II, III and IV respectively). The phases of enrolment are described in more detail in the cohort profile paper and its update.^1,2^ The total sample size for analyses using any data collected after the age of seven is therefore 15,447 pregnancies, resulting in 15,658 foetuses. Of these 14,901 children were alive at 1 year of age.

### ALSPAC non-response weights

Response at each timepoint was modelled based on measures collected during the perinatal period, with missing data imputed using multivariable imputation by chained equations.^3^ Weighting variables were: maternal age at delivery, education, socioeconomic status, ethnicity, household characteristics, smoking during pregnancy and pre- and post-natal depression scores. For the second timepoint and beyond, the non-response weight from the previous timepoint was included as an additional predictor. To maintain consistency with GUS, where non-response weights were provided alongside each data release, we used the inverse predicted probability of responding (after rescaling to the available sample size) to weight ALSPAC data at each timepoint.^4^

### Statistical analyses: additional details

All statistical analyses were performed in R version 4.5.0.^5^ Age-only, unadjusted and fully-adjusted trajectory models were fit using the “lme4” package version 1.1-37.^6^ Participant age was re-centred to the sample mean in each cohort (ALSPAC 11.19 years; GUS 8.78 years) to facilitate model fitting. Mean-centred age was then back-transformed to aid interpretability of figures and estimated mean SDQ scores by age. Model parameters were estimated using Maximum Likelihood Estimation and Bound Optimization by Quadratic Approximation (“bobyqa”), with a set maximum of 200,000 iterations. Models which did not converge or had singular fit were excluded from comparisons. p-values and 95% confidence intervals were calculated using the “lmerTest” package version 3.1-3,^7^ via Satterthwaite’s method of approximating degrees of freedom in mixed-effect models.

Marginal means of predicted SDQ scores were estimated using the “emmeans” package version 1.11.1,^8^ averaging over all categorical covariate levels (child sex, household income, area deprivation) and calculated at the mean maternal mental health score for each cohort. Estimated marginal means were weighted proportional to the frequency of each combination of covariate levels in the original sample.

### Additional results: FI trajectories in ALSPAC

*Conduct problems:* Conduct problem scores followed a cubic trajectory in ALSPAC (Figure S1, Table S3). Prior to adjusting for covariates, both High and Low FI groups had higher conduct problem trajectories compared to the No FI group (Figure S3, Table S5). In fully-adjusted models, High and Low FI remained associated with higher (worse) trajectories compared to No FI, though differences in model intercept reduced in magnitude (Table S9). In terms of individual trajectories, all FI groups showed an initial steady decrease in conduct problems which plateaued in early-mid adolescence (Figure 1A). Conduct problems then decreased from age 15 onward in both High and No FI trajectories; the Low FI trajectory showed a slight increase between ages 11-15 followed by a sharper decline (Figure 1A, Table 2).

*Emotional symptoms:* Emotional symptom scores followed a cubic trajectory in ALSPAC (Figure S1, Table S3). Both High and Low FI groups had higher trajectories compared to the No FI group in unadjusted models (Figure S3, Table S6). After adjusting for covariates, High FI remained associated with higher emotional symptoms compared to No FI; the association for Low FI was attenuated (Table S10). Trajectories of emotional symptoms differed in shape between FI groups. In the No FI group, emotional symptoms remained relatively stable until around age 16, where scores began to gradually increase (Figure 1B, Table 2). For the Low FI group, this increase in emotional symptoms began earlier – around age 11 – and scores continued to rise at a steady pace throughout adolescence. The High FI trajectory, in contrast, had higher initial emotional symptoms which increased throughout childhood (ages 7-9). Emotional problems then declined between ages 9-15; this was followed by a much steeper increase in emotional problems compared to No and Low FI trajectories (Figure 1B, Table 2).

*Hyperactivity/inattention:* Hyperactivity/inattention scores followed a quartic trajectory in ALSPAC (Figure S1, Table S3). Prior to adjusting for covariates, both High and Low FI groups had higher conduct problem trajectories compared to the No FI group (Figure S3, Table S7). In fully-adjusted models, High and Low FI remained associated with higher (worse) trajectories compared to No FI, though differences in model intercept reduced in magnitude (Table S11). In terms of trajectory shape, all FI groups followed a similar pattern. In all groups, hyperactivity/inattention scores decreased from late childhood until early adolescence (Figure 1C, Table 2). Scores then increased slightly from age 11-15 before sharply decreasing once more; this was more pronounced for Low and High FI trajectories (Figure 1C, Table 2).

*Peer problems:* Peer problem scores followed a quartic trajectory in ALSPAC (Figure S1, Table S3). Prior to adjusting for covariates, both High and Low FI groups had higher trajectories compared to the No FI group (Figure S3, Table S8). In fully-adjusted models, High and Low FI remained associated with higher levels of peer problems compared to No FI, though differences in model intercept reduced in magnitude (Table S12). In terms of individual trajectories, Low and No FI trajectories were similar in shape. Peer problems gradually increased from childhood to mid-adolescence in both groups, reaching a peak around age 15 (Figure 1D). For the High FI group, peer problem scores increased more sharply in late childhood and were maintained over adolescence (Figure 1D, Table 2).

### Additional results: FI trajectories in GUS

*Conduct problems:* Conduct problems followed a cubic trajectory in GUS (Figure S2, Table S4). Prior to adjusting for covariates, High and Low FI were both associated with higher (worse) conduct problem trajectories compared to No FI (Figure S4, Table S13). High FI remained associated with higher overall trajectories in the fully-adjusted model, however associations for Low FI were attenuated (Table S17). In terms of individual trajectories, Low and No FI groups had overlapping trajectories (Figure 1E, Table 4); all groups had similarly shaped trajectories. Conduct problems decreased more rapidly in late childhood compared to adolescence and at a similar rate across FI levels (Figure 1E, Table 2).

*Emotional symptoms:* Emotional symptoms followed a cubic trajectory in GUS (Figure S2, Table S4). In the unadjusted model, High and Low FI were both associated with higher emotional problem trajectories compared to No FI (Figure S4, Table S14). After adjusting for covariates, High FI remained associated with worse emotional symptom trajectories; the association for Low FI was attenuated (Table S14). All FI groups showed a steady increase in emotional symptoms from childhood to mid-adolescence (age 5-13); this was slightly steeper for Low vs No FI trajectories (Figure 1F, Table 2). For Low and No FI groups, this increase slowed around age 13 and scores began to decline after age 14. This slowing was less pronounced for the High FI trajectory where emotional symptoms continued to gradually rise after age 13 (Figure 1F, Table 2).

*Hyperactivity/inattention:* Hyperactivity/inattention scores followed a linear trajectory in GUS (Figure S2, Table S4). Prior to adjusting for covariates, both High and Low FI were associated with higher trajectories of hyperactivity/inattention compared to No FI (Figure S4, Table S15). High FI remained associated with higher levels of hyperactivity/inattention after adjusting for covariates; the association for Low FI was attenuated (Table S19). In terms of individual trajectories, all FI groups showed a similar linear decrease in hyperactivity/inattention over childhood and adolescence. This was marginally slower in the High FI group compared to Low/No FI groups (Figure 1G, Table 2 and 4).

*Peer problems:* Peer problems followed a quartic trajectory in GUS (Figure S2, Table S4). Prior to adjusting for covariates, both High and Low FI groups had higher trajectories compared to the No FI group (Figure S4, Table S16). In fully-adjusted models, High and Low FI remained associated with higher levels of peer problems compared to No FI, though differences in model intercept reduced in magnitude (Table S20). In terms of individual trajectories, the No FI group showed a steady increase in peer problems from age 5-15, at an approximately linear pace (Figure 1H). While all groups started with similar levels of peer problems, the increase in peer problem scores was more pronounced for High and Low FI groups, which had similar levels of peer problems at age 15 (Figure 1H, Table 4). Instead of an approximately linear increase, both High and Low FI trajectories showed a steeper initial rise in peer problems which slowed slightly in early adolescence before sharply increasing again from around age 13/14 (Figure 1H).

### Figure S1: Age-only trajectory models in ALSPAC

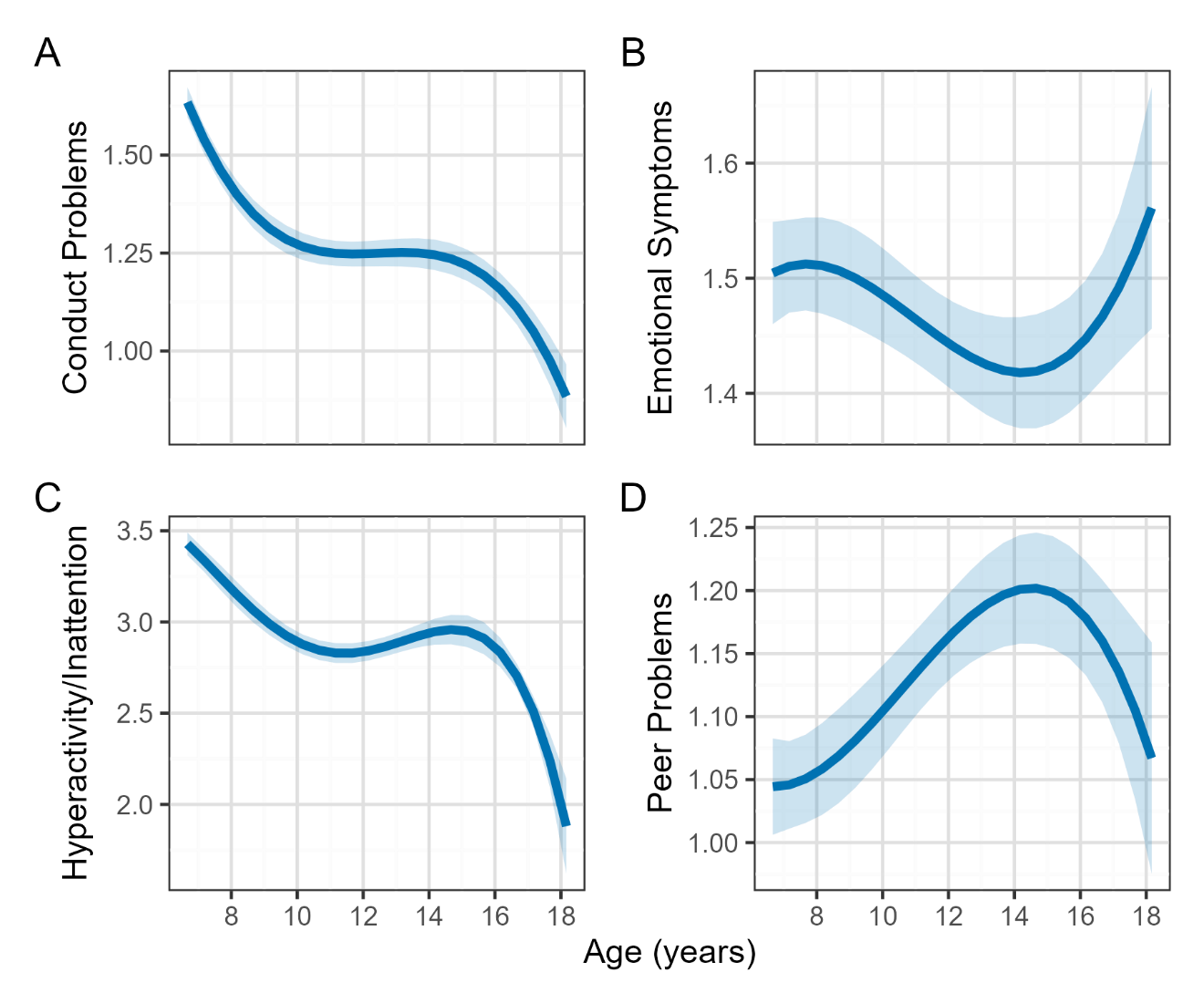

Best-fitting age-only trajectory models of SDQ subscale scores in ALSPAC. Conduct problems (A), emotional symptoms (B) and peer problems (D) followed a cubic trajectory model; hyperactivity/inattention scores (C) followed a quartic trajectory model.

ALSPAC = Avon Longitudinal Study of Parents and Children; SDQ = Strengths and Difficulties Questionnaire.

### Figure S2: Age-only trajectory models in GUS

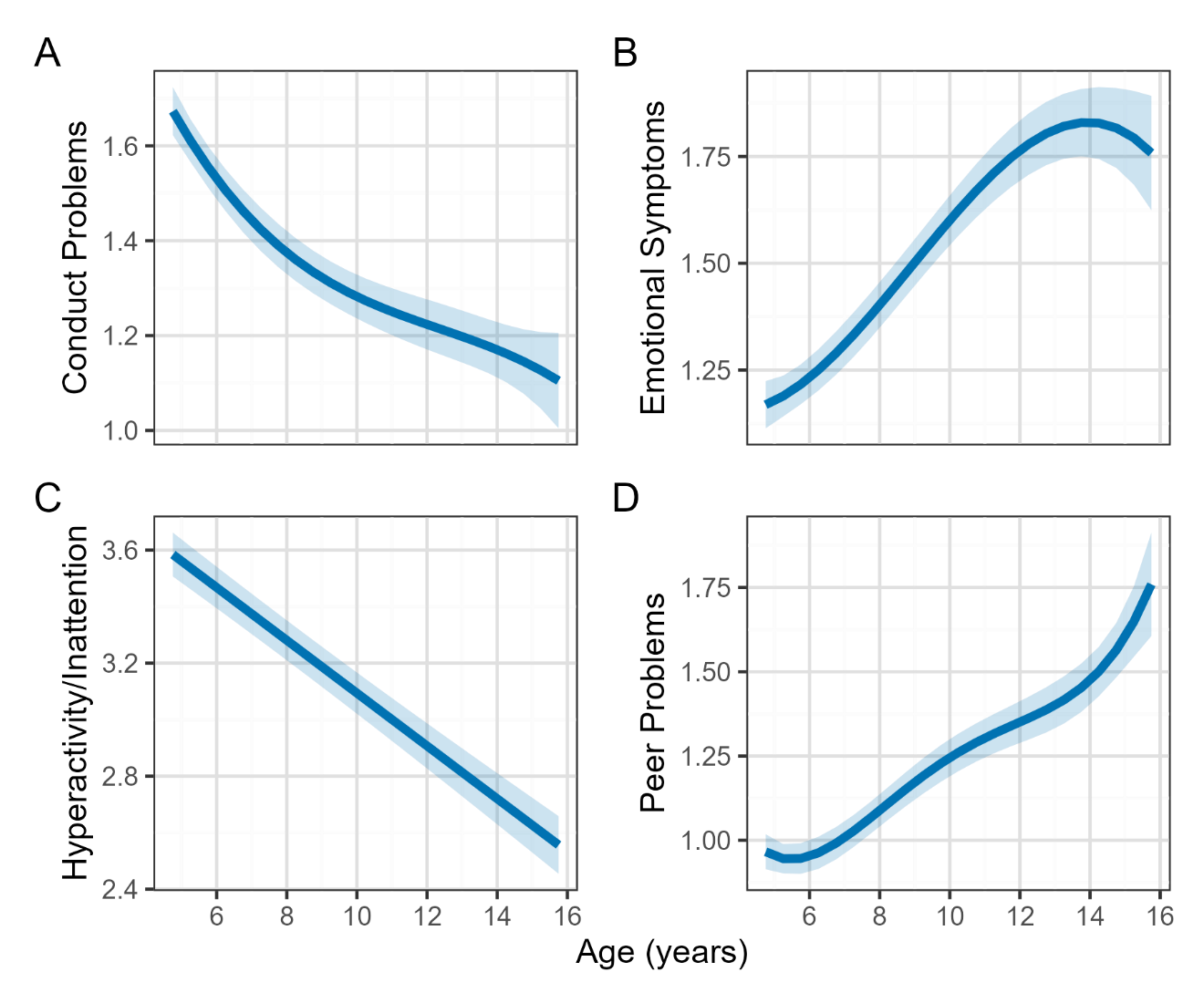

Best-fitting age-only trajectory models of SDQ subscale scores in GUS. Conduct problems (A) and emotional symptoms (B) followed a cubic trajectory model; hyperactivity/inattention (C) scores followed a linear trajectory model; peer problems (D) followed a quartic trajectory model.

GUS = Growing Up in Scotland; SDQ = Strengths and Difficulties Questionnaire.

### Figure S3: Unadjusted trajectory models by FI level in ALSPAC

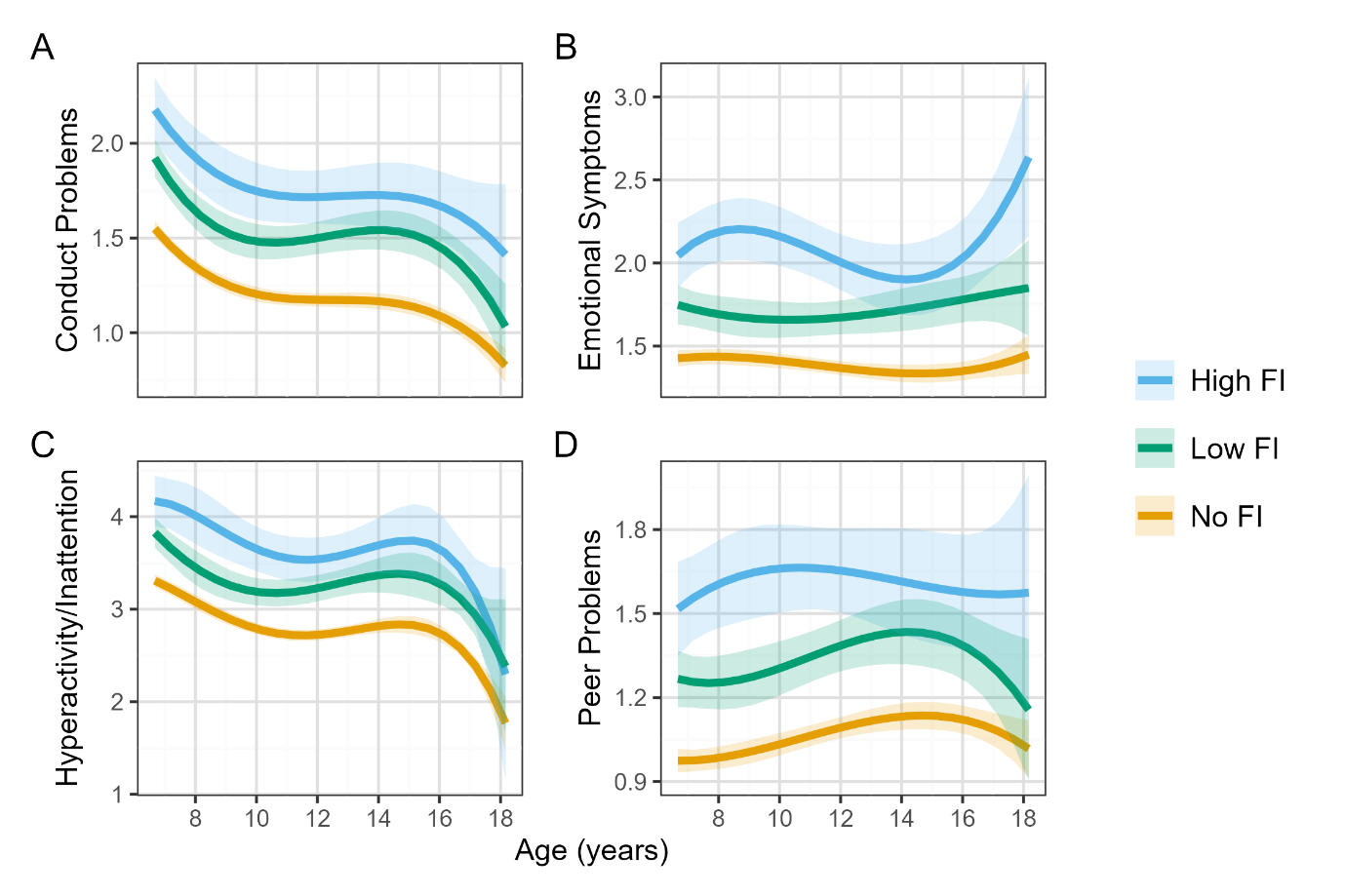

Unadjusted trajectories of SDQ subscale scores in ALSPAC, stratified by food insecurity exposure at age 5.

ALSPAC = Avon Longitudinal Study of Parents and Children; FI = food insecurity; SDQ = Strengths and Difficulties Questionnaire.

### Figure S4: Unadjusted trajectory models by FI level in GUS

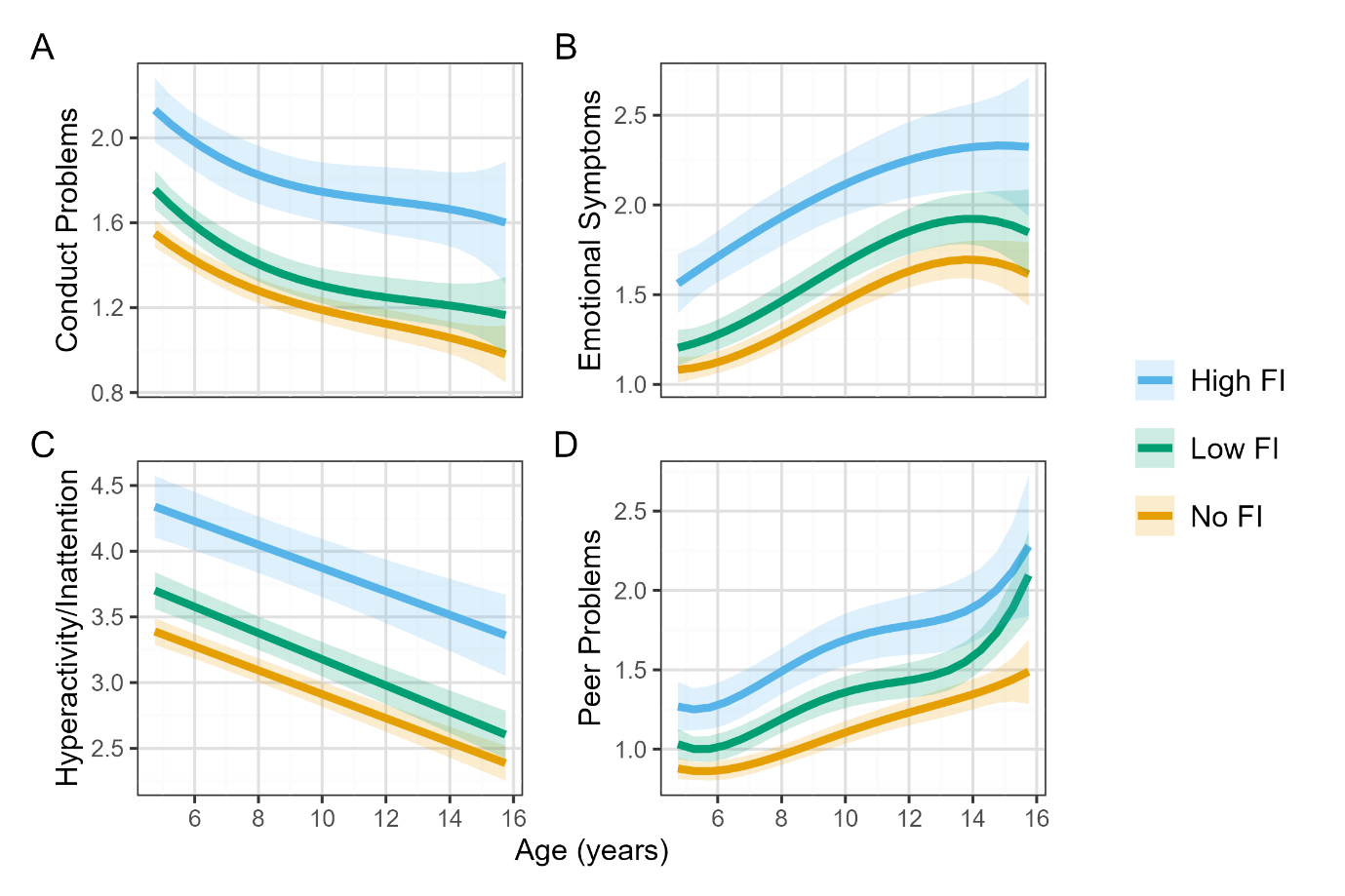

Unadjusted trajectories of SDQ subscale scores in GUS, stratified by food insecurity exposure at age 5.

GUS = Growing Up in Scotland; FI = food insecurity; SDQ = Strengths and Difficulties Questionnaire.

### Table S1: ALSPAC sample demographics

|  | | **Included (n = 6182)** | **Excluded (n = 8847)** |
| --- | --- | --- | --- |
| *Baseline demographics ^a^* | | | |
| Sex | Male | 3151 (50.97%) | 4532 (51.23%) |
|  | Female | 3031 (49.03%) | 4315 (48.77%) |
| Maternal age at delivery | | 28.86 (4.52) | 27.04 (5.93) |
| Housing tenure | Mortgage/Owned | 4947 (83.92%) | 4915 (69.13%) |
|  | Private rented | 325 (5.51%) | 653 (9.18%) |
|  | Subsidised rented | 623 (10.57%) | 1542 (21.69%) |
| Parity | 1st | 2816 (46.63%) | 3048 (43.17%) |
|  | 2nd | 2189 (36.25%) | 2386 (33.79%) |
|  | 3rd + | 1034 (17.12%) | 1627 (23.04%) |
| Smoking in first trimester | No | 4853 (79.52%) | 5119 (70.78%) |
|  | Yes | 1250 (20.48%) | 2113 (29.22%) |
| Maternal education | >=A level | 2568 (42.33%) | 1830 (28.58%) |
|  | <O level | 1313 (21.65%) | 2440 (38.11%) |
|  | O level | 2185 (36.02%) | 2133 (33.31%) |
| Occupational class ^b^ | I-II | 2156 (41.2%) | 1617 (33.23%) |
|  | III-V | 3077 (58.8%) | 3249 (66.77%) |
| Child ethnicity | White | 5757 (96.32%) | 5759 (93.63%) |
|  | Non-white ^c^ | 220 (3.68%) | 392 (6.37%) |
| Financial difficulties | No | 4549 (76.85%) | 4431 (71.24%) |
|  | Yes | 1370 (23.15%) | 1789 (28.76%) |
| EPDS | Prenatal | 6.67 (4.89) | 7.49 (5.24) |
|  | Postnatal | 5.82 (4.59) | 6.31 (4.97) |
| *At child age ~5 years* | | | |
| Difficulty affording food ^d^ | Not difficult | 4972 (80.43%) | 2195 (78.03%) |
|  | Low difficult | 881 (14.25%) | 450 (16%) |
|  | Fairly difficult | 282 (4.56%) | 131 (4.66%) |
|  | Very difficult | 47 (0.76%) | 37 (1.32%) |
| FI level | No FI | 4972 (80.43%) | 2195 (78.03%) |
|  | Low FI | 881 (14.25%) | 450 (16%) |
|  | High FI | 329 (5.32%) | 168 (5.97%) |
| Weekly household income | >£400 | 1560 (25.23%) | 551 (20.92%) |
|  | £300 to £299 | 1403 (22.69%) | 470 (17.84%) |
|  | £200 to £299 | 1770 (28.63%) | 734 (27.87%) |
|  | £100 to £199 | 1022 (16.53%) | 537 (20.39%) |
|  | <£100 | 427 (6.91%) | 342 (12.98%) |
| EPDS covariate | | 8.09 (3.86) | 8.37 (4.07) |
| IMD Quintile | Least deprived | 2046 (33.1%) | 558 (27.17%) |
|  | 2 | 1548 (25.04%) | 441 (21.47%) |
|  | 3 | 1071 (17.32%) | 357 (17.38%) |
|  | 5 | 957 (15.48%) | 380 (18.5%) |
|  | Most deprived | 560 (9.06%) | 318 (15.48%) |

Continued on next page.

### Table S1: ALSPAC sample demographics (continued)

|  |  | **Included (n = 6182)** | **Excluded (n = 8847)** |
| --- | --- | --- | --- |
| *Longitudinal measures* | | | |
| Age at sweep | Sweep KQ | 6.78 (0.1) | 6.81 (0.13) |
|  | Sweep KU | 9.64 (0.12) | 9.67 (0.15) |
|  | Sweep KW | 11.71 (0.12) | 11.73 (0.16) |
|  | Sweep TA | 13.15 (0.18) | 13.17 (0.2) |
|  | Sweep TC | 16.81 (0.35) | 16.91 (0.38) |
| Sample size at sweep | Sweep KQ | 5760 (93.17%) | 2634 (27.83%) |
|  | Sweep KU | 5293 (85.62%) | 2903 (30.68%) |
|  | Sweep KW | 4912 (79.46%) | 2543 (26.87%) |
|  | Sweep TA | 4730 (76.51%) | 2406 (25.43%) |
|  | Sweep TC | 3894 (62.99%) | 1809 (19.12%) |

Demographic characteristics of ALSPAC participants included and excluded from analyses. ALSPAC = Avon Longitudinal Study of Parents and Children; EPDS = Edinburgh Postnatal Depression Scale; FI = food insecurity; IMD = Index of Multiple Deprivation.

^a^ Perinatal variables measured between 1990-1993, used for calculating non-response weights.

^b^ Occupational class in ALSPAC was derived using the UK Registrar General’s occupational coding (SOC 90).

^c^ Child ethnicity was derived from mother reports of her and her partner’s ethnic backgrounds. Child ethnicity was coded as non-white if either parent had a non-white background, which included: Black Caribbean, Black African, Other black, Indian, Pakistani, Bangladeshi, Chinese, Other.

^d^ Used to derive FI level

### Table S2: GUS sample demographics

|  | | **Included (n = 3167)** | **Excluded (n = 666)** |
| --- | --- | --- | --- |
| *Baseline demographics* | | | |
| Sex | Male | 1633 (51.56%) | 323 (48.5%) |
|  | Female | 1534 (48.44%) | 343 (51.5%) |
| Maternal age at delivery | <20 | 129 (4.08%) | 52 (7.83%) |
|  | 20-29 | 1112 (35.2%) | 287 (43.22%) |
|  | 30-39 | 1797 (56.89%) | 303 (45.63%) |
|  | 40+ | 121 (3.83%) | 22 (3.31%) |
| Housing tenure | Mortgage/owned | 2306 (74.85%) | 402 (61.85%) |
|  | Private rented | 157 (5.1%) | 43 (6.62%) |
|  | Subsidised rented | 618 (20.06%) | 205 (31.54%) |
| Birth order | 1st born | 1529 (48.28%) | 338 (50.75%) |
|  | 2nd born | 1079 (34.07%) | 218 (32.73%) |
|  | 3rd+ born | 559 (17.65%) | 110 (16.52%) |
| Smoking in pregnancy | No | 2492 (79.92%) | 484 (74.69%) |
|  | Yes | 626 (20.08%) | 164 (25.31%) |
| Maternal education | <GCSE | 194 (6.13%) | 79 (11.92%) |
|  | >=A level | 2147 (67.84%) | 378 (57.01%) |
|  | GCSE | 824 (26.03%) | 206 (31.07%) |
| NS-SEC Occupational class | Managerial, administrative & professional | 1358 (42.89%) | 195 (29.28%) |
|  | Intermediate –Unemployed | 1808 (57.11%) | 471 (70.72%) |
| Child ethnicity | White | 3072 (97.06%) | 621 (93.24%) |
|  | Non-white ^a^ | 93 (2.94%) | 45 (6.76%) |
| *At child age ~5 years* | | | |
| Cost of food affects what you give child to eat ^b^ | Not at all | 1877 (59.27%) | 399 (59.91%) |
|  | A little | 956 (30.19%) | 196 (29.43%) |
|  | A fair amount | 258 (8.15%) | 54 (8.11%) |
|  | A lot | 76 (2.4%) | 17 (2.55%) |
| FI level | No FI | 1877 (59.27%) | 399 (59.91%) |
|  | Low FI | 956 (30.19%) | 196 (29.43%) |
|  | High FI | 334 (10.55%) | 71 (10.66%) |
| Annual household income ^c^ | <£12,217 | 583 (18.41%) | 138 (31.36%) |
|  | >=£12,217 <£19,643 | 698 (22.04%) | 103 (23.41%) |
|  | >=£19,643 < £29,126 | 585 (18.47%) | 59 (13.41%) |
|  | >=£29,126 < £37,857 | 768 (24.25%) | 84 (19.09%) |
|  | >=£37,857 | 533 (16.83%) | 56 (12.73%) |
| SF-12 MCS covariate | | 50.6 (9.07) | 49.95 (10.22) |

Continued on next page.

### Table S2: GUS sample demographics (continued)

|  |  | **Included (n = 3167)** | **Excluded (n = 666)** |
| --- | --- | --- | --- |
| *At child age ~5 years* | | | |
| SIMD quintile | Least deprived | 744 (23.49%) | 105 (15.77%) |
|  | 2 | 714 (22.54%) | 137 (20.57%) |
|  | 3 | 645 (20.37%) | 120 (18.02%) |
|  | 4 | 543 (17.15%) | 151 (22.67%) |
|  | Most deprived | 521 (16.45%) | 153 (22.97%) |
| *Longitudinal measures* | | | |
| Age at sweep | Sweep 5 | 4.85 (0.04) | 4.86 (0.04) |
|  | Sweep 6 | 5.85 (0.04) | 5.86 (0.04) |
|  | Sweep 7 | 7.87 (0.06) | 7.88 (0.07) |
|  | Sweep 8 | 10.18 (0.3) | 10.17 (0.32) |
|  | Sweep 9 | 12.56 (0.31) | 12.55 (0.33) |
|  | Sweep 10 | 14.64 (0.39) | 14.62 (0.41) |
| Sample size at sweep | Sweep 5 | 3167 (100%) | 666 (100%) |
|  | Sweep 6 | 3167 (100%) | 369 (55.41%) |
|  | Sweep 7 | 2935 (92.67%) | 395 (59.31%) |
|  | Sweep 8 | 2699 (85.22%) | 350 (52.55%) |
|  | Sweep 9 | 2525 (79.73%) | 308 (46.25%) |
|  | Sweep 10 | 2330 (73.57%) | 262 (39.34%) |

Demographic characteristics of GUS participants included and excluded from analyses. GUS = Growing Up in Scotland; NS-SEC = National Statistics Socio-economic Classification; FI = food insecurity; SF-12 MCS = Short-Form 12 health survey Mental Component Scale; SIMD = Scottish Index of Multiple Deprivation.

^a^ Non-white ethnicity includes the following parent-reported backgrounds: Mixed - White and Black Caribbean, Mixed - White and Black African, Mixed - White and Asian, Any other mixed background, Asian/Asian British – Indian, Asian/Asian British – Pakistani, Asian/Asian British – Bangladeshi, Any other Asian/Asian British background, Black/Black British – Caribbean, Black/Black British – African, Any other Black/Black British background, Chinese, Any other non-white background.

^b^ Used to derive FI level

^c^ Equivalised to account for household composition

### Table S3: ALSPAC age-only model comparisons

| **SDQ subscale** | **Model** | **AIC** | **BIC** | **Deviance** | **χ^2^** |
| --- | --- | --- | --- | --- | --- |
| Conduct problems | Linear | 77244.28 | 77292.85 | 77232.28 |  |
|  | Quadratic | 77168.6 | 77225.27 | 77154.6 | 77.67 *** |
|  | **Cubic** | 77096.5 | 77161.26 | 77080.5 | 74.11 *** |
|  | Quartic | 77098.5 | 77171.36 | 77080.5 | 0.001 |
| Emotional symptoms | Linear | 87920.69 | 87969.26 | 87908.69 |  |
|  | Quadratic | 87918.19 | 87974.86 | 87904.19 | 4.50 * |
|  | **Cubic** | 87915.52 | 87980.29 | 87899.52 | 4.66 * |
|  | Quartic | 87917.47 | 87990.33 | 87899.47 | 0.06 |
| Hyperactivity/Inattention | Linear | 95365.31 | 95413.89 | 95353.31 |  |
|  | Quadratic | 95271.17 | 95327.84 | 95257.17 | 96.15 *** |
|  | Cubic | 95168.66 | 95233.43 | 95152.66 | 104.51 *** |
|  | **Quartic** | 95158.45 | 95231.32 | 95140.45 | 12.21 *** |
| Peer problems | Linear | 81284.97 | 81333.54 | 81272.97 |  |
|  | Quadratic | 81274.68 | 81331.35 | 81260.68 | 12.29 *** |
|  | **Cubic** | 81270.8 | 81335.56 | 81254.8 | 5.88 * |
|  | Quartic | 81270.58 | 81343.45 | 81252.58 | 2.21 |

Model fit indices and likelihood ratio tests comparing age-only trajectory models with increasing-order polynomial terms. The best-fitting trajectory for each SDQ subscale is highlighted in bold. Smaller values for AIC, BIC and deviance indicate better model fit.

ALSPAC = Avon Longitudinal Study of Parents and Children; SDQ = Strengths and Difficulties Questionnaire; AIC = Akaike Information Criterion; BIC = Bayesian Information Criterion.

**p*<.05; ****p*<.001

### Table S4: GUS age-only model comparisons

| **SDQ subscale** | **Model** | **AIC** | **BIC** | **Deviance** | **χ^2^** |
| --- | --- | --- | --- | --- | --- |
| Conduct problems | Linear | 52425.15 | 52471.32 | 52413.15 |  |
|  | Quadratic | 52391.19 | 52445.06 | 52377.19 | 35.96 *** |
|  | **Cubic** | 52389.21 | 52450.78 | 52373.21 | 3.98 * |
|  | Quartic | 52389.51 | 52458.77 | 52371.51 | 1.70 |
| Emotional symptoms | Linear | 59485.85 | 59532.02 | 59473.85 |  |
|  | Quadratic | 59479.36 | 59533.23 | 59465.36 | 8.49 ** |
|  | **Cubic** | 59469.66 | 59531.22 | 59453.66 | 11.71 *** |
|  | Quartic | 59471.62 | 59540.87 | 59453.62 | 0.04 |
| Hyperactivity/Inattention | **Linear** | 65061.06 | 65107.23 | 65049.06 |  |
|  | Quadratic | 65061.18 | 65115.05 | 65047.18 | 1.88 |
|  | Cubic | 65061.55 | 65123.11 | 65045.55 | 1.63 |
|  | Quartic | 65062.56 | 65131.82 | 65044.56 | 0.99 |
| Peer problems | Linear | 54880.69 | 54926.86 | 54868.69 |  |
|  | Quadratic | 54877.44 | 54931.3 | 54863.44 | 5.25 * |
|  | Cubic | 54878.16 | 54939.72 | 54862.16 | 1.28 |
|  | **Quartic** | 54871.73 | 54940.98 | 54853.73 | 8.43 ** |

Model fit indices and likelihood ratio tests comparing age-only trajectory models with increasing-order polynomial terms. The best-fitting trajectory for each SDQ subscale is highlighted in bold. Smaller values for AIC, BIC and deviance indicate better model fit.

GUS = Growing Up in Scotland; SDQ = Strengths and Difficulties Questionnaire; AIC = Akaike Information Criterion; BIC = Bayesian Information Criterion.

**p*<.05; ***p*<.01; ****p*<.001

### Table S5: Conduct problem trajectories in ALSPAC – unadjusted model

| *Predictors* | *Estimates* | *CI* | *p* |
| --- | --- | --- | --- |
| (Intercept) | 1.18 | 1.14 – 1.21 | **<0.001** |
| Age* | -0.01 | -0.02 – 0.00 | 0.119 |
| FI [High] | 0.54 | 0.40 – 0.68 | **<0.001** |
| FI [Low] | 0.30 | 0.21 – 0.39 | **<0.001** |
| Age^2 | 0.01 | 0.01 – 0.01 | **<0.001** |
| Age^3 | -0.00 | -0.00 – -0.00 | **<0.001** |
| Age × FI [High] | 0.00 | -0.05 – 0.05 | 0.960 |
| Age × FI [Low] | 0.03 | -0.01 – 0.06 | 0.113 |
| FI [High] × Age^2 | 0.00 | -0.00 – 0.01 | 0.315 |
| FI [Low] × Age^2 | 0.00 | -0.00 – 0.01 | 0.077 |
| FI [High] × Age^3 | -0.00 | -0.00 – 0.00 | 0.762 |
| FI [Low] × Age^3 | -0.00 | -0.00 – 0.00 | 0.062 |
| **Random Effects** | | | |
| σ^2^ | 0.67 | | |
| τ_00_ _ID_ | 1.13 | | |
| τ_11_ _ID.Age_ | 0.01 | | |
| ρ_01_ _ID_ | -0.14 | | |
| ICC | 0.65 | | |
| N _ID_ | 6182 | | |
| Observations | 24243 | | |
| Marginal R^2^ / Conditional R^2^ | 0.029 / 0.658 | | |

*mean-centred to 11.19 years

### Table S6: Emotional symptom trajectories in ALSPAC – unadjusted model

| *Predictors* | *Estimates* | *CI* | *p* |
| --- | --- | --- | --- |
| (Intercept) | 1.39 | 1.34 – 1.43 | **<0.001** |
| Age* | -0.02 | -0.04 – -0.01 | **0.006** |
| FI [High] | 0.69 | 0.52 – 0.86 | **<0.001** |
| FI [Low] | 0.28 | 0.17 – 0.38 | **<0.001** |
| Age^2 | -0.00 | -0.00 – 0.00 | 0.960 |
| Age^3 | 0.00 | -0.00 – 0.00 | 0.059 |
| Age × FI [High] | -0.06 | -0.13 – 0.01 | 0.077 |
| Age × FI [Low] | 0.03 | -0.01 – 0.07 | 0.149 |
| FI [High] × Age^2 | -0.00 | -0.01 – 0.01 | 0.503 |
| FI [Low] × Age^2 | 0.00 | -0.00 – 0.01 | 0.053 |
| FI [High] × Age^3 | 0.00 | 0.00 – 0.01 | **0.031** |
| FI [Low] × Age^3 | -0.00 | -0.00 – 0.00 | 0.292 |
| **Random Effects** | | | |
| σ^2^ | 1.13 | | |
| τ_00_ _ID_ | 1.44 | | |
| τ_11_ _ID.age.cent_ | 0.01 | | |
| ρ_01_ _ID_ | 0.03 | | |
| ICC | 0.58 | | |
| N _ID_ | 6182 | | |
| Observations | 24243 | | |
| Marginal R^2^ / Conditional R^2^ | 0.012 / 0.589 | | |

*mean-centred to 11.19 years

### Table S7: Hyperactivity/inattention trajectories in ALSPAC – unadjusted model

| *Predictors* | *Estimates* | *CI* | *p* |
| --- | --- | --- | --- |
| (Intercept) | 2.72 | 2.66 – 2.78 | **<0.001** |
| Age* | -0.02 | -0.04 – -0.00 | **0.035** |
| FI [High] | 0.82 | 0.58 – 1.07 | **<0.001** |
| FI [Low] | 0.46 | 0.31 – 0.62 | **<0.001** |
| Age^2 | 0.03 | 0.02 – 0.04 | **<0.001** |
| Age^3 | -0.00 | -0.00 – -0.00 | **0.001** |
| Age^4 | -0.00 | -0.00 – -0.00 | **0.001** |
| Age × FI [High] | -0.02 | -0.10 – 0.06 | 0.627 |
| Age × FI [Low] | 0.05 | 0.00 – 0.10 | **0.032** |
| FI [High] × Age^2 | 0.01 | -0.03 – 0.06 | 0.551 |
| FI [Low] × Age^2 | -0.00 | -0.03 – 0.03 | 0.793 |
| FI [High] × Age^3 | 0.00 | -0.00 – 0.01 | 0.588 |
| FI [Low] × Age^3 | -0.00 | -0.01 – 0.00 | 0.078 |
| FI [High] × Age^4 | -0.00 | -0.00 – 0.00 | 0.546 |
| FI [Low] × Age^4 | 0.00 | -0.00 – 0.00 | 0.542 |
| **Random Effects** | | | |
| σ^2^ | 1.26 | | |
| τ_00_ _ID_ | 3.24 | | |
| τ_11_ _ID.age.cent_ | 0.03 | | |
| ρ_01_ _ID_ | -0.22 | | |
| ICC | 0.74 | | |
| N _ID_ | 6182 | | |
| Observations | 24243 | | |
| Marginal R^2^ / Conditional R^2^ | 0.026 / 0.744 | | |

*mean-centred to 11.19 years

### Table S8: Peer problem trajectories in ALSPAC – unadjusted model

| *Predictors* | *Estimates* | *CI* | *p* |
| --- | --- | --- | --- |
| (Intercept) | 1.07 | 1.03 – 1.11 | **<0.001** |
| Age* | 0.03 | 0.02 – 0.04 | **<0.001** |
| FI [High] | 0.59 | 0.44 – 0.74 | **<0.001** |
| FI [Low] | 0.28 | 0.19 – 0.38 | **<0.001** |
| Age^2 | -0.00 | -0.00 – 0.00 | 0.275 |
| Age^3 | -0.00 | -0.00 – -0.00 | **0.034** |
| Age × FI [High] | -0.04 | -0.10 – 0.02 | 0.218 |
| Age × FI [Low] | 0.01 | -0.03 – 0.05 | 0.542 |
| FI [High] × Age^2 | -0.00 | -0.01 – 0.00 | 0.183 |
| FI [Low] × Age^2 | -0.00 | -0.00 – 0.00 | 0.999 |
| FI [High] × Age^3 | 0.00 | -0.00 – 0.00 | 0.297 |
| FI [Low] × Age^3 | -0.00 | -0.00 – 0.00 | 0.408 |
| **Random Effects** | | | |
| σ^2^ | 0.87 | | |
| τ_00_ _ID_ | 1.18 | | |
| τ_11_ _ID.age.cent_ | 0.01 | | |
| ρ_01_ _ID_ | 0.15 | | |
| ICC | 0.59 | | |
| N _ID_ | 6182 | | |
| Observations | 24243 | | |
| Marginal R^2^ / Conditional R^2^ | 0.011 / 0.598 | | |

*mean-centred to 11.19 years

### Table S9: Conduct problem trajectories in ALSPAC – fully-adjusted model

| *Predictors* | *Estimates* | *CI* | *p* |
| --- | --- | --- | --- |
| (Intercept) | 0.77 | 0.67 – 0.86 | **<0.001** |
| Age* | -0.01 | -0.02 – 0.00 | 0.141 |
| FI [High] | 0.24 | 0.09 – 0.38 | **0.001** |
| FI [Low] | 0.14 | 0.05 – 0.23 | **0.003** |
| Age^2 | 0.01 | 0.01 – 0.01 | **<0.001** |
| Age^3 | -0.00 | -0.00 – -0.00 | **<0.001** |
| sex [Female] | -0.15 | -0.20 – -0.09 | **<0.001** |
| income300-399 | -0.03 | -0.11 – 0.05 | 0.497 |
| income200-299 | 0.04 | -0.04 – 0.12 | 0.271 |
| income100-199 | 0.17 | 0.07 – 0.27 | **0.001** |
| income [<100] | 0.35 | 0.22 – 0.48 | **<0.001** |
| epds | 0.05 | 0.05 – 0.06 | **<0.001** |
| IMD [2] | -0.03 | -0.11 – 0.04 | 0.421 |
| IMD [3] | 0.00 | -0.08 – 0.09 | 0.928 |
| IMD [4] | 0.11 | 0.02 – 0.20 | **0.019** |
| IMD [5] | 0.16 | 0.05 – 0.28 | **0.006** |
| Age × FI [High] | 0.00 | -0.05 – 0.05 | 0.948 |
| Age × FI [Low] | 0.03 | -0.01 – 0.06 | 0.121 |
| FI [High] × Age^2 | 0.00 | -0.00 – 0.01 | 0.294 |
| FI [Low] × Age^2 | 0.00 | -0.00 – 0.01 | 0.067 |
| FI [High] × Age^3 | -0.00 | -0.00 – 0.00 | 0.746 |
| FI [Low] × Age^3 | -0.00 | -0.00 – 0.00 | 0.062 |
| **Random Effects** | | | |
| σ^2^ | 0.67 | | |
| τ_00_ _ID_ | 1.06 | | |
| τ_11_ _ID.age.cent_ | 0.01 | | |
| ρ_01_ _ID_ | -0.13 | | |
| ICC | 0.64 | | |
| N _ID_ | 6182 | | |
| Observations | 24243 | | |
| Marginal R^2^ / Conditional R^2^ | 0.062 / 0.658 | | |

*mean-centred to 11.19 years

### Table S10: Emotional symptom trajectories in ALSPAC – fully-adjusted model

| *Predictors* | *Estimates* | *CI* | *p* |
| --- | --- | --- | --- |
| (Intercept) | 0.56 | 0.46 – 0.67 | **<0.001** |
| Age* | -0.02 | -0.04 – -0.01 | **0.006** |
| FI [High] | 0.37 | 0.20 – 0.54 | **<0.001** |
| FI [Low] | 0.09 | -0.01 – 0.20 | 0.087 |
| Age^2 | -0.00 | -0.00 – 0.00 | 0.942 |
| Age^3 | 0.00 | -0.00 – 0.00 | 0.051 |
| sex [Female] | 0.27 | 0.21 – 0.34 | **<0.001** |
| income300-399 | 0.10 | 0.01 – 0.20 | **0.034** |
| income200-299 | 0.18 | 0.09 – 0.27 | **<0.001** |
| income100-199 | 0.25 | 0.14 – 0.37 | **<0.001** |
| income [<100] | 0.23 | 0.08 – 0.39 | **0.003** |
| epds | 0.08 | 0.07 – 0.09 | **<0.001** |
| IMD [2] | -0.02 | -0.11 – 0.06 | 0.584 |
| IMD [3] | -0.03 | -0.13 – 0.07 | 0.595 |
| IMD [4] | -0.06 | -0.16 – 0.05 | 0.269 |
| IMD [5] | -0.04 | -0.17 – 0.10 | 0.580 |
| Age × FI [High] | -0.06 | -0.13 – 0.01 | 0.082 |
| Age × FI [Low] | 0.03 | -0.01 – 0.07 | 0.143 |
| FI [High] × Age^2 | -0.00 | -0.01 – 0.01 | 0.529 |
| FI [Low] × Age^2 | 0.01 | 0.00 – 0.01 | **0.039** |
| FI [High] × Age^3 | 0.00 | 0.00 – 0.01 | **0.036** |
| FI [Low] × Age^3 | -0.00 | -0.00 – 0.00 | 0.262 |
| **Random Effects** | | | |
| σ^2^ | 1.13 | | |
| τ_00_ _ID_ | 1.32 | | |
| τ_11_ _ID.age.cent_ | 0.01 | | |
| ρ_01_ _ID_ | 0.02 | | |
| ICC | 0.56 | | |
| N _ID_ | 6182 | | |
| Observations | 24243 | | |
| Marginal R^2^ / Conditional R^2^ | 0.052 / 0.587 | | |

*mean-centred to 11.19 years

### Table S11: Hyperactivity/inattention trajectories in ALSPAC – fully-adjusted model

| *Predictors* | *Estimates* | *CI* | *p* |
| --- | --- | --- | --- |
| (Intercept) | 2.30 | 2.15 – 2.45 | **<0.001** |
| Age* | -0.02 | -0.04 – -0.00 | **0.047** |
| FI [High] | 0.35 | 0.10 – 0.59 | **0.005** |
| FI [Low] | 0.19 | 0.03 – 0.35 | **0.016** |
| Age^2 | 0.03 | 0.02 – 0.04 | **<0.001** |
| Age^3 | -0.00 | -0.00 – -0.00 | **0.001** |
| Age^4 | -0.00 | -0.00 – -0.00 | **0.001** |
| sex [Female] | -0.84 | -0.93 – -0.75 | **<0.001** |
| income300-399 | 0.10 | -0.03 – 0.23 | 0.136 |
| income200-299 | 0.22 | 0.09 – 0.35 | **0.001** |
| income100-199 | 0.46 | 0.30 – 0.61 | **<0.001** |
| income [<100] | 0.61 | 0.40 – 0.83 | **<0.001** |
| epds | 0.08 | 0.07 – 0.10 | **<0.001** |
| IMD [2] | -0.01 | -0.13 – 0.12 | 0.929 |
| IMD [3] | -0.02 | -0.16 – 0.11 | 0.733 |
| IMD [4] | 0.10 | -0.05 – 0.25 | 0.193 |
| IMD [5] | 0.08 | -0.11 – 0.26 | 0.419 |
| Age × FI [High] | -0.02 | -0.10 – 0.06 | 0.606 |
| Age × FI [Low] | 0.05 | 0.00 – 0.10 | **0.039** |
| FI [High] × Age^2 | 0.01 | -0.03 – 0.06 | 0.549 |
| FI [Low] × Age^2 | -0.00 | -0.03 – 0.03 | 0.819 |
| FI [High] × Age^3 | 0.00 | -0.00 – 0.01 | 0.573 |
| FI [Low] × Age^3 | -0.00 | -0.01 – 0.00 | 0.086 |
| FI [High] × Age^4 | -0.00 | -0.00 – 0.00 | 0.547 |
| FI [Low] × Age^4 | 0.00 | -0.00 – 0.00 | 0.564 |
| **Random Effects** | | | |
| σ^2^ | 1.26 | | |
| τ_00_ _ID_ | 2.92 | | |
| τ_11_ _ID.age.cent_ | 0.03 | | |
| ρ_01_ _ID_ | -0.21 | | |
| ICC | 0.72 | | |
| N _ID_ | 6182 | | |
| Observations | 24243 | | |
| Marginal R^2^ / Conditional R^2^ | 0.090 / 0.744 | | |

*mean-centred to 11.19 years

### Table S12: Peer problem trajectories in ALSPAC – fully-adjusted model

| *Predictors* | *Estimates* | *CI* | *p* |
| --- | --- | --- | --- |
| (Intercept) | 0.70 | 0.61 – 0.80 | **<0.001** |
| Age* | 0.03 | 0.02 – 0.04 | **<0.001** |
| FI [High] | 0.30 | 0.14 – 0.45 | **<0.001** |
| FI [Low] | 0.12 | 0.02 – 0.21 | **0.021** |
| Age^2 | -0.00 | -0.00 – 0.00 | 0.259 |
| Age^3 | -0.00 | -0.00 – -0.00 | **0.037** |
| sex [Female] | -0.20 | -0.26 – -0.14 | **<0.001** |
| income300-399 | -0.01 | -0.10 – 0.08 | 0.817 |
| income200-299 | 0.13 | 0.05 – 0.21 | **0.002** |
| income100-199 | 0.30 | 0.20 – 0.40 | **<0.001** |
| income [<100] | 0.40 | 0.26 – 0.54 | **<0.001** |
| epds | 0.05 | 0.04 – 0.06 | **<0.001** |
| IMD [2] | -0.03 | -0.11 – 0.04 | 0.389 |
| IMD [3] | 0.02 | -0.07 – 0.10 | 0.725 |
| IMD [4] | 0.02 | -0.08 – 0.11 | 0.719 |
| IMD [5] | 0.03 | -0.09 – 0.15 | 0.578 |
| Age × FI [High] | -0.04 | -0.09 – 0.02 | 0.225 |
| Age × FI [Low] | 0.01 | -0.03 – 0.05 | 0.557 |
| FI [High] × Age^2 | -0.00 | -0.01 – 0.00 | 0.197 |
| FI [Low] × Age^2 | 0.00 | -0.00 – 0.00 | 0.957 |
| FI [High] × Age^3 | 0.00 | -0.00 – 0.00 | 0.310 |
| FI [Low] × Age^3 | -0.00 | -0.00 – 0.00 | 0.407 |
| **Random Effects** | | | |
| σ^2^ | 0.87 | | |
| τ_00_ _ID_ | 1.11 | | |
| τ_11_ _ID.age.cent_ | 0.01 | | |
| ρ_01_ _ID_ | 0.17 | | |
| ICC | 0.58 | | |
| N _ID_ | 6182 | | |
| Observations | 24243 | | |
| Marginal R^2^ / Conditional R^2^ | 0.040 / 0.597 | | |

*mean-centred to 11.19 years

### Table S13: Conduct problem trajectories in GUS – unadjusted model

| *Predictors* | *Estimates* | *CI* | *p* |
| --- | --- | --- | --- |
| (Intercept) | 1.24 | 1.18 – 1.30 | **<0.001** |
| Age* | -0.05 | -0.06 – -0.03 | **<0.001** |
| FI [High] | 0.55 | 0.40 – 0.69 | **<0.001** |
| FI [Low] | 0.12 | 0.02 – 0.22 | **0.020** |
| Age^2 | 0.01 | 0.00 – 0.01 | **0.001** |
| Age^3 | -0.00 | -0.00 – 0.00 | 0.176 |
| Age × FI [High] | 0.00 | -0.03 – 0.04 | 0.804 |
| Age × FI [Low] | -0.01 | -0.03 – 0.02 | 0.620 |
| FI [High] × Age^2 | 0.00 | -0.00 – 0.01 | 0.516 |
| FI [Low] × Age^2 | 0.00 | -0.00 – 0.01 | 0.208 |
| FI [High] × Age^3 | -0.00 | -0.00 – 0.00 | 0.812 |
| FI [Low] × Age^3 | -0.00 | -0.00 – 0.00 | 0.854 |
| **Random Effects** | | | |
| σ^2^ | 0.81 | | |
| τ_00_ _Idnumber_ | 1.11 | | |
| τ_11_ _Idnumber.age.cent_ | 0.02 | | |
| ρ_01_ _Idnumber_ | 0.09 | | |
| ICC | 0.62 | | |
| N _Idnumber_ | 3167 | | |
| Observations | 16239 | | |
| Marginal R^2^ / Conditional R^2^ | 0.029 / 0.627 | | |

*mean-centred to 8.78 years

### Table S14: Emotional symptom trajectories in GUS – unadjusted model

| *Predictors* | *Estimates* | *CI* | *p* |
| --- | --- | --- | --- |
| (Intercept) | 1.35 | 1.28 – 1.42 | **<0.001** |
| Age* | 0.10 | 0.08 – 0.12 | **<0.001** |
| FI [High] | 0.66 | 0.48 – 0.84 | **<0.001** |
| FI [Low] | 0.20 | 0.08 – 0.32 | **0.001** |
| Age^2 | 0.00 | -0.00 – 0.01 | 0.359 |
| Age^3 | -0.00 | -0.00 – -0.00 | **0.004** |
| Age × FI [High] | -0.00 | -0.05 – 0.05 | 0.918 |
| Age × FI [Low] | 0.01 | -0.02 – 0.05 | 0.532 |
| FI [High] × Age^2 | -0.01 | -0.02 – 0.00 | 0.133 |
| FI [Low] × Age^2 | -0.00 | -0.01 – 0.00 | 0.636 |
| FI [High] × Age^3 | 0.00 | -0.00 – 0.00 | 0.322 |
| FI [Low] × Age^3 | 0.00 | -0.00 – 0.00 | 0.916 |
| **Random Effects** | | | |
| σ^2^ | 1.29 | | |
| τ_00_ _Idnumber_ | 1.53 | | |
| τ_11_ _Idnumber.age.cent_ | 0.03 | | |
| ρ_01_ _Idnumber_ | 0.48 | | |
| ICC | 0.59 | | |
| N _Idnumber_ | 3167 | | |
| Observations | 16239 | | |
| Marginal R^2^ / Conditional R^2^ | 0.030 / 0.606 | | |

*mean-centred to 8.78 years

### Table S15: Hyperactivity/inattention trajectories in GUS – unadjusted model

| *Predictors* | *Estimates* | *CI* | *p* |
| --- | --- | --- | --- |
| (Intercept) | 3.02 | 2.93 – 3.11 | **<0.001** |
| Age* | -0.09 | -0.10 – -0.08 | **<0.001** |
| FI [High] | 0.96 | 0.72 – 1.19 | **<0.001** |
| FI [Low] | 0.28 | 0.12 – 0.43 | **0.001** |
| Age × FI [High] | 0.00 | -0.03 – 0.03 | 0.896 |
| Age × FI [Low] | -0.01 | -0.03 – 0.01 | 0.428 |
| **Random Effects** | | | |
| σ^2^ | 1.59 | | |
| τ_00_ _Idnumber_ | 3.63 | | |
| τ_11_ _Idnumber.age.cent_ | 0.04 | | |
| ρ_01_ _Idnumber_ | 0.06 | | |
| ICC | 0.72 | | |
| N _Idnumber_ | 3167 | | |
| Observations | 16239 | | |
| Marginal R^2^ / Conditional R^2^ | 0.032 / 0.727 | | |

*mean-centred to 8.78 years

### Table S16: Peer problem trajectories in GUS – unadjusted model

| *Predictors* | *Estimates* | *CI* | *p* |
| --- | --- | --- | --- |
| (Intercept) | 1.02 | 0.95 – 1.08 | **<0.001** |
| Age* | 0.07 | 0.05 – 0.10 | **<0.001** |
| FI [High] | 0.56 | 0.40 – 0.73 | **<0.001** |
| FI [Low] | 0.25 | 0.14 – 0.36 | **<0.001** |
| Age^2 | 0.00 | -0.01 – 0.01 | 0.807 |
| Age^3 | -0.00 | -0.00 – 0.00 | 0.073 |
| Age^4 | 0.00 | -0.00 – 0.00 | 0.311 |
| Age × FI [High] | 0.03 | -0.02 – 0.09 | 0.219 |
| Age × FI [Low] | 0.02 | -0.02 – 0.06 | 0.331 |
| FI [High] × Age^2 | -0.01 | -0.03 – 0.00 | 0.154 |
| FI [Low] × Age^2 | -0.01 | -0.03 – -0.00 | **0.049** |
| FI [High] × Age^3 | -0.00 | -0.00 – 0.00 | 0.570 |
| FI [Low] × Age^3 | -0.00 | -0.00 – 0.00 | 0.496 |
| FI [High] × Age^4 | 0.00 | -0.00 – 0.00 | 0.300 |
| FI [Low] × Age^4 | 0.00 | -0.00 – 0.00 | 0.085 |
| **Random Effects** | | | |
| σ^2^ | 0.96 | | |
| τ_00_ _Idnumber_ | 1.26 | | |
| τ_11_ _Idnumber.age.cent_ | 0.02 | | |
| ρ_01_ _Idnumber_ | 0.45 | | |
| ICC | 0.61 | | |
| N _Idnumber_ | 3167 | | |
| Observations | 16239 | | |
| Marginal R^2^ / Conditional R^2^ | 0.029 / 0.620 | | |

*mean-centred to 8.78 years

### Table S17: Conduct problem trajectories in GUS – fully-adjusted model

| *Predictors* | *Estimates* | *CI* | *p* |
| --- | --- | --- | --- |
| (Intercept) | 2.56 | 2.30 – 2.81 | **<0.001** |
| Age* | -0.05 | -0.06 – -0.03 | **<0.001** |
| FI [High] | 0.21 | 0.07 – 0.36 | **0.004** |
| FI [Low] | 0.01 | -0.09 – 0.11 | 0.851 |
| Age^2 | 0.01 | 0.00 – 0.01 | **<0.001** |
| Age^3 | -0.00 | -0.00 – 0.00 | 0.153 |
| Sex [F] | -0.23 | -0.30 – -0.15 | **<0.001** |
| EqvIncome [1] | 0.51 | 0.37 – 0.65 | **<0.001** |
| EqvIncome [2] | 0.16 | 0.03 – 0.29 | **0.017** |
| EqvIncome [3] | 0.10 | -0.03 – 0.23 | 0.135 |
| EqvIncome [4] | 0.10 | -0.02 – 0.22 | 0.115 |
| SF12-MCS | -0.03 | -0.03 – -0.02 | **<0.001** |
| SIMD [2] | 0.05 | -0.06 – 0.17 | 0.343 |
| SIMD [3] | 0.18 | 0.06 – 0.29 | **0.004** |
| SIMD [4] | 0.15 | 0.03 – 0.28 | **0.018** |
| SIMD [5] | 0.28 | 0.15 – 0.42 | **<0.001** |
| Age × FI [High] | 0.00 | -0.03 – 0.04 | 0.803 |
| Age × FI [Low] | -0.01 | -0.03 – 0.02 | 0.593 |
| FI [High] × Age^2 | 0.00 | -0.00 – 0.01 | 0.532 |
| FI [Low] × Age^2 | 0.00 | -0.00 – 0.01 | 0.207 |
| FI [High] × Age^3 | -0.00 | -0.00 – 0.00 | 0.819 |
| FI [Low] × Age^3 | -0.00 | -0.00 – 0.00 | 0.870 |
| **Random Effects** | | | |
| σ^2^ | 0.81 | | |
| τ_00_ _Idnumber_ | 0.98 | | |
| τ_11_ _Idnumber.age.cent_ | 0.02 | | |
| ρ_01_ _Idnumber_ | 0.13 | | |
| ICC | 0.59 | | |
| N _Idnumber_ | 3167 | | |
| Observations | 16239 | | |
| Marginal R^2^ / Conditional R^2^ | 0.093 / 0.627 | | |

*mean-centred to 8.78 years

### Table S18: Emotional symptom trajectories in GUS – fully-adjusted model

| *Predictors* | *Estimates* | *CI* | *p* |
| --- | --- | --- | --- |
| (Intercept) | 2.94 | 2.65 – 3.23 | **<0.001** |
| Age* | 0.10 | 0.08 – 0.12 | **<0.001** |
| FI [High] | 0.38 | 0.20 – 0.55 | **<0.001** |
| FI [Low] | 0.10 | -0.02 – 0.22 | 0.091 |
| Age^2 | 0.00 | -0.00 – 0.01 | 0.352 |
| Age^3 | -0.00 | -0.00 – -0.00 | **0.003** |
| Sex [F] | 0.04 | -0.05 – 0.12 | 0.373 |
| EqvIncome [1] | 0.18 | 0.02 – 0.34 | **0.027** |
| EqvIncome [2] | 0.11 | -0.04 – 0.26 | 0.153 |
| EqvIncome [3] | -0.00 | -0.15 – 0.15 | 0.964 |
| EqvIncome [4] | -0.11 | -0.25 – 0.03 | 0.116 |
| SF12-MCS | -0.03 | -0.04 – -0.03 | **<0.001** |
| SIMD [2] | 0.08 | -0.05 – 0.21 | 0.234 |
| SIMD [3] | 0.07 | -0.06 – 0.21 | 0.283 |
| SIMD [4] | 0.17 | 0.02 – 0.31 | **0.021** |
| SIMD [5] | 0.21 | 0.06 – 0.35 | **0.007** |
| Age × FI [High] | -0.00 | -0.05 – 0.05 | 0.961 |
| Age × FI [High] | 0.01 | -0.02 – 0.05 | 0.513 |
| FI [High] × Age^2 | -0.01 | -0.02 – 0.00 | 0.135 |
| FI [Low] × Age^2 | -0.00 | -0.01 – 0.00 | 0.664 |
| FI [High] × Age^3 | 0.00 | -0.00 – 0.00 | 0.323 |
| FI [Low] × Age^3 | 0.00 | -0.00 – 0.00 | 0.930 |
| **Random Effects** | | | |
| σ^2^ | 1.29 | | |
| τ_00_ _Idnumber_ | 1.38 | | |
| τ_11_ _Idnumber.age.cent_ | 0.03 | | |
| ρ_01_ _Idnumber_ | 0.47 | | |
| ICC | 0.57 | | |
| N _Idnumber_ | 3167 | | |
| Observations | 16239 | | |
| Marginal R^2^ / Conditional R^2^ | 0.065 / 0.600 | | |

*mean-centred to 8.78 years

### Table S19: Hyperactivity/inattention trajectories in GUS – fully-adjusted model

| *Predictors* | *Estimates* | *CI* | *p* |
| --- | --- | --- | --- |
| (Intercept) | 5.19 | 4.75 – 5.63 | **<0.001** |
| Age* | -0.09 | -0.10 – -0.08 | **<0.001** |
| FI [High] | 0.41 | 0.18 – 0.64 | **<0.001** |
| FI [Low] | 0.10 | -0.05 – 0.25 | 0.183 |
| Sex [F] | -0.88 | -1.01 – -0.75 | **<0.001** |
| EqvIncome [1] | 0.76 | 0.52 – 1.01 | **<0.001** |
| EqvIncome [2] | 0.35 | 0.12 – 0.57 | **0.003** |
| EqvIncome [3] | 0.18 | -0.05 – 0.41 | 0.124 |
| EqvIncome [4] | -0.02 | -0.23 – 0.19 | 0.855 |
| SF12-MCS | -0.04 | -0.05 – -0.03 | **<0.001** |
| SIMD [2] | 0.07 | -0.12 – 0.27 | 0.458 |
| SIMD [3] | 0.19 | -0.02 – 0.39 | 0.073 |
| SIMD [4] | 0.22 | -0.00 – 0.44 | 0.050 |
| SIMD [5] | 0.53 | 0.30 – 0.76 | **<0.001** |
| Age × FI [High] | 0.00 | -0.03 – 0.03 | 0.893 |
| Age × FI [Low] | -0.01 | -0.03 – 0.01 | 0.396 |
| **Random Effects** | | | |
| σ^2^ | 1.59 | | |
| τ_00_ _Idnumber_ | 3.12 | | |
| τ_11_ _Idnumber.age.cent_ | 0.04 | | |
| ρ_01_ _Idnumber_ | 0.05 | | |
| ICC | 0.69 | | |
| N _Idnumber_ | 3167 | | |
| Observations | 16239 | | |
| Marginal R^2^ / Conditional R^2^ | 0.119 / 0.727 | | |

*mean-centred to 8.78 years

### Table S20: Peer problem trajectories in GUS – fully-adjusted model

| *Predictors* | *Estimates* | *CI* | *p* |
| --- | --- | --- | --- |
| (Intercept) | 2.04 | 1.78 – 2.29 | **<0.001** |
| Age* | 0.07 | 0.05 – 0.10 | **<0.001** |
| FI [High] | 0.24 | 0.08 – 0.41 | **0.004** |
| FI [Low] | 0.14 | 0.03 – 0.25 | **0.011** |
| Age^2 | 0.00 | -0.01 – 0.01 | 0.834 |
| Age^3 | -0.00 | -0.00 – 0.00 | 0.062 |
| Age^4 | 0.00 | -0.00 – 0.00 | 0.287 |
| Sex [F] | -0.20 | -0.28 – -0.13 | **<0.001** |
| EqvIncome [1] | 0.48 | 0.34 – 0.62 | **<0.001** |
| EqvIncome [2] | 0.24 | 0.11 – 0.38 | **<0.001** |
| EqvIncome [3] | 0.21 | 0.08 – 0.35 | **0.002** |
| EqvIncome [4] | -0.01 | -0.13 – 0.12 | 0.900 |
| SF12-MCS | -0.02 | -0.03 – -0.02 | **<0.001** |
| SIMD [2] | 0.09 | -0.03 – 0.21 | 0.130 |
| SIMD [3] | 0.07 | -0.05 – 0.19 | 0.283 |
| SIMD [4] | 0.24 | 0.12 – 0.37 | **<0.001** |
| SIMD [5] | 0.32 | 0.18 – 0.45 | **<0.001** |
| Age × FI [High] | 0.04 | -0.02 – 0.09 | 0.206 |
| Age × FI [Low] | 0.02 | -0.02 – 0.06 | 0.346 |
| FI [High] × Age^2 | -0.01 | -0.03 – 0.00 | 0.147 |
| FI [Low] × Age^2 | -0.01 | -0.02 – 0.00 | 0.053 |
| FI [High] × Age^3 | -0.00 | -0.00 – 0.00 | 0.561 |
| FI [Low] × Age^3 | -0.00 | -0.00 – 0.00 | 0.511 |
| FI [High] × Age^4 | 0.00 | -0.00 – 0.00 | 0.289 |
| FI [Low] × Age^4 | 0.00 | -0.00 – 0.00 | 0.091 |
| **Random Effects** | | | |
| σ^2^ | 0.96 | | |
| τ_00_ _Idnumber_ | 1.12 | | |
| τ_11_ _Idnumber.age.cent_ | 0.02 | | |
| ρ_01_ _Idnumber_ | 0.45 | | |
| ICC | 0.58 | | |
| N _Idnumber_ | 3167 | | |
| Observations | 16239 | | |
| Marginal R^2^ / Conditional R^2^ | 0.075 / 0.616 | | |

*mean-centred to 8.78 years

4	Lubian K, Tipping S, Bradshaw P. Survey weights and longitudinal analysis. .

5	R Core Team. R: A Language and Environment for Statistical Computing. 2025. https://www.R-project.org/.

6	Bates D, Mächler M, Bolker B, Walker S. Fitting Linear Mixed-Effects Models Using lme4. *Journal of Statistical Software* 2015; **67**: 1–48.

7	Kuznetsova A, Brockhoff PB, Christensen RHB. lmerTest Package: Tests in Linear Mixed Effects Models. *Journal of Statistical Software* 2017; **82**: 1–26.

8	Lenth RV. emmeans: Estimated Marginal Means, aka Least-Squares Means. 2025. DOI:10.32614/CRAN.package.emmeans.
